# Precision Diagnostics: Using Islet Autoantibodies to Characterize Heterogeneity in Type 1 Diabetes

**DOI:** 10.1101/2023.04.18.23288756

**Authors:** Jamie L. Felton, Maria J. Redondo, Richard A. Oram, Cate Speake, S. Alice Long, Suna Onengut-Gumuscu, Stephen S. Rich, Gabriela SF Monaco, Arianna Harris-Kawano, Dianna Perez, Zeb Saeed, Benjamin Hoag, Rashmi Jain, Carmella Evans-Molina, Linda A. DiMeglio, Heba Ismail, Dana Dabelea, Randi K. Johnson, Marzhan Urazbayeva, John M. Wentworth, Kurt J. Griffin, Emily K. Sims, the ADA/EASD Precision Medicine in Diabetes Initiative Consortium

## Abstract

**Background:** Heterogeneity exists in type 1 diabetes (T1D) development and presentation. Islet autoantibodies form the foundation for T1D diagnostic and staging efforts. We hypothesized that autoantibodies can be used to identify heterogeneity in T1D before, at, and after diagnosis, and in response to disease modifying therapies. at clinically relevant timepoints throughout T1D progression.

**Methods:** We performed a systematic review assessing 10 years of original research studies examining relationships between autoantibodies and heterogeneity during disease progression, at the time of diagnosis, after diagnosis, and in response to disease modifying therapies in individuals at risk for T1D or within 1 year of T1D diagnosis.

**Results:** 10,067 papers were screened. Out of 151 that met data extraction criteria, 90 studies characterized heterogeneity before clinical diagnosis. Autoantibody type/target was most commonly examined, followed by autoantibody number, titer, order of seroconversion, affinity, and novel islet autoantibodies/epitopes. Recurring themes included positive relationships of autoantibody number and specific types and titers with disease progression, differing clinical phenotypes based on the order of autoantibody seroconversion, and interactions with age and genetics. Overall, reporting of autoantibody assay performance was commonly included; however, only 43% (65/151) included information about autoantibody assay standardization efforts. Populations studied were almost exclusively of European ancestry.

**Conclusions:** Current evidence most strongly supports the application of autoantibody features to more precisely define T1D before clinical diagnosis. Our findings support continued use of pre-clinical staging paradigms based on autoantibody number and suggest that additional autoantibody features, particularly when considered in relation to age and genetic risk, could offer more precise stratification. Increased participation in autoantibody standardization efforts is a critical step to improving future applicability of autoantibody-based precision medicine in T1D.

**Plain Language Summary:** We performed a systematic review to ascertain whether islet autoantibodies, biomarkers of autoimmunity against insulin-producing cells, could aid in stratifying individuals with different clinical presentations of type 1 diabetes. We found existing evidence most strongly supporting the application of these biomarkers to the period before clinical diagnosis, when certain autoantibody features (number, type) and the age when they develop, can provide important information for patients and care providers on what to expect for future type 1 diabetes progression.

## Introduction

Type 1 diabetes (T1D) results from the immune-mediated destruction of insulin-producing pancreatic beta cells (1). Clinical disease is characterized by progressive hyperglycemia that, if left untreated, leads to ketoacidosis and death. T1D can be managed with exogenous insulin, and while technology surrounding glucose monitoring and insulin delivery have revolutionized diabetes care, effective disease management remains difficult, time-consuming, and costly. Islet autoantibodies that recognize insulin (IAA), glutamic acid decarboxylase (GAD), protein phosphatase-like IA-2 (IA-2), zinc transporter 8 (ZnT8), and islet cell cytoplasmic antigen (ICA), are well-validated predictors of risk and disease progression and have been proposed as diagnostic markers of presymptomatic stages of T1D. Stage 1 T1D is defined by the presence of multiple islet autoantibodies and normal glucose tolerance. This progresses to stage 2 T1D (multiple islet autoantibodies and dysglycemia) and ultimately stage 3 T1D with onset of clinical symptoms, typically requiring treatment with exogenous insulin (2). Understanding the pathophysiology that drives T1D progression through these stages remains critical to developing interventions to pause or reverse disease progression. However, vast heterogeneity exists in T1D progression, presentation, and responses to interventions. These differences suggest that differences in clinical features or presentation of disease progression and response to treatment could reflect discreet pathophysiological mechanisms. Along these lines, if distinct etiologic mechanisms are responsible for different forms of disease, it may be that specific subsets of individuals with T1D will respond better to specific disease-modifying therapies with improved risk/benefit ratios. Therefore, precision approaches to diagnosis may be necessary to effect disease modification in T1D.

The *Precision Medicine in Diabetes Initiative* (PMDI) was established in 2018 by the American Diabetes Association (ADA) in partnership with the European Association for the Study of Diabetes (EASD). The ADA/EASD PMDI includes global thought leaders in precision diabetes medicine who are working to address the burgeoning need for better diabetes prevention and care through precision medicine (3). This Systematic Review is written on behalf of the ADA/EASD PMDI as part of a comprehensive evidence evaluation in support of the 2^nd^ International Consensus Report on Precision Diabetes Medicine.

Multiple studies and clinical trials have investigated the impact of genetics, immune markers, metabolic function, and environmental factors, on the development and progression of T1D (4). Some of these works have identified subgroups of individuals who may derive greater benefit than others from particular therapies. In this systematic review, we sought to identify aspects of precision medicine that have the potential to be adopted into clinical practice over the next 10 years. Given the substantial body of work focused on optimization, reproducibility, and validation of islet autoantibodies as biomarkers of islet autoimmunity (5–8), and their increased use in clinical practice since the development of the T1D staging system (2), we chose to focus on islet autoantibodies as an individual feature of disease. We hypothesized that islet autoantibodies can be used to identify unique phenotypes of disease progression and presentation at four clinically-relevant timepoints: prior to clinical (stage 3) T1D diagnosis, at stage 3 T1D onset, after stage 3 T1D diagnosis, and in response to disease modifying therapy at diagnosis (new onset trials) or before the time of stage 3 T1D diagnosis (prevention trials).

## Methods

### Data Source

We developed a search strategy using an iterative process that involved identification of Medical Subject Headings (MeSH) and text words, followed by refinement based on a sensitivity check for key articles identified by group members. On 10/25/21 “Precision Medicine”[Mesh] AND (Latent Autoimmune Diabetes in Adults [Mesh] OR “Diabetes Mellitus, Type 1 “[Mesh]) was applied as an initial search strategy to PubMed. Based on identification of only 128 papers (which were mostly reviews), the search strategy was expanded to include additional terms linked to precision medicine (Supplemental Fig. 1). This strategy was applied to PubMed and EMBASE databases by Lund University librarians on 6/14/2022.

### Study Selection

The Covidence platform was used for stages of systematic review. To be included, studies must have involved individuals with high genetic risk (based on family history or genotype), single islet autoantibody positivity, stage 1 T1D (multiple islet autoantibody positivity and normal glucose tolerance), stage 2 T1D (positive islet autoantibodies and abnormal glucose tolerance), or stage 3 T1D (overt hyperglycemia, clinical symptoms of untreated T1D). Individuals with stage 3 T1D must have been within one year of diagnosis.

Eligible study types included randomized controlled trials; systematic reviews or meta-analyses of randomized controlled trials; cross-sectional studies; open-label extension studies; prospective observational studies; retrospective observational studies; and post-hoc analyses. Studies must have had a total sample size ≥ 10/group studied and been published as a full paper in English in a peer-reviewed journal within 10 years of the search (2011-2022). Studies of non-T1D populations, unclearly classified diabetes populations, or mixed populations that included T1D among other diabetes types (type 2 diabetes, Latent Autoimmune Diabetes in Adults (LADA), gestational diabetes, or hypothetical cohort) were excluded. Several key articles identified by the group of experts that also met inclusion criteria but were not included in the search results because of search restrictions made to improve search feasibility, were also included in the analysis.

Investigators independently screened and reviewed each potentially relevant article according to preliminary eligibility criteria determined by members of the review team. For Level 1 screening two investigators per article screened each title and abstract. Discordant assessments were discussed and resolved by consensus or arbitration after consultation with a member of the review leadership team (JLF, RO, KJG, MJR, or EKS). In January of 2022, to improve review feasibility, the decision was made to limit articles at the Level 2 screening step using additional inclusion criteria. Here articles were further limited to exposures testing detection of abnormal islet autoantibodies (i.e., presence, total number, type, or titer) and addressing outcomes related to progression to multiple antibody positivity or diabetes, heterogenous presentation of disease, progression of C-peptide loss after diabetes develops, or response to treatment. For Level 2 screening of eligible articles, full texts were retrieved and reviewed by two independent reviewers using the inclusion/exclusion criteria. Discordant assessments were similarly discussed and resolved.

### Data Extraction

Two independent investigators from the writing group extracted data from each article meeting inclusion criteria, with consensus determined by a member of the leadership team. Extracted data included details on participant characteristics, intervention outcomes, methods, and conclusions of precision analyses on disease progression or treatment response. Investigators performed quality assessments using a modified version of the Joanna Briggs Institute’s critical appraisal checklist tool for cohort studies (https://jbi.global/critical-appraisal-tools) in tandem for each eligible study to determine overall risk of bias.

### Data Analysis and Synthesis

Because of heterogeneity of included studies (i.e., design, population, exposure, and outcomes tested) we were unable to perform a meta-analysis. Instead, we provide completed summaries of relevant studies (Supplemental Tables 1-4). The protocol of this review was registered at Prospero.com prior to implementation (available at https://www.crd.york.ac.uk/prospero/display_record.php?ID=CRD42022340047)

## Results

### Literature search and screening results

Of the 10,067 papers identified via the literature search of PubMed and Embase databases, 151 met inclusion criteria for extraction (Figure 1). We categorized studies based on clinically relevant timepoints: 90 characterized differences in rates of progression and clinical features prior to stage 3 T1D and were categorized as “prior to diagnosis”; 44 assessed differences in metabolic or immune features at stage 3 T1D onset and were categorized as “at diagnosis”; 11 characterized metabolic decline after diagnosis and were categorized as “after diagnosis”; and 13 assessed differences in responses to disease-modifying therapies tested in clinical trials and were categorized as “treatment response” (Table 1). Of note, some papers included multiple studies of several outcomes; therefore, a total of 158 studies were identified from 151 papers.

**Table 1.** Classification of literature search results by clinically relevant timepoint.

| **Clinical Timepoint** | **Description** | **Number of studies identified** |
| --- | --- | --- |
| **Prior to diagnosis** | Studies that assessed differences in rates of progression and clinical features during the period leading up to stage 3 T1D diagnosis. | 90 |
| **At diagnosis** | Studies that assessed heterogeneity in clinical features at the time of stage 3 T1D diagnosis. | 44 |
| **After diagnosis** | Studies that characterized metabolic decline (and preservation of endogenous insulin production) after stage 3 T1D diagnosis. | 11 |
| **Treatment response** | Studies that assessed heterogeneity in responses to disease modifying therapies tested in clinical trials in subjects at or before stage 3 T1D diagnosis. | 13 |

**Figure 1.**
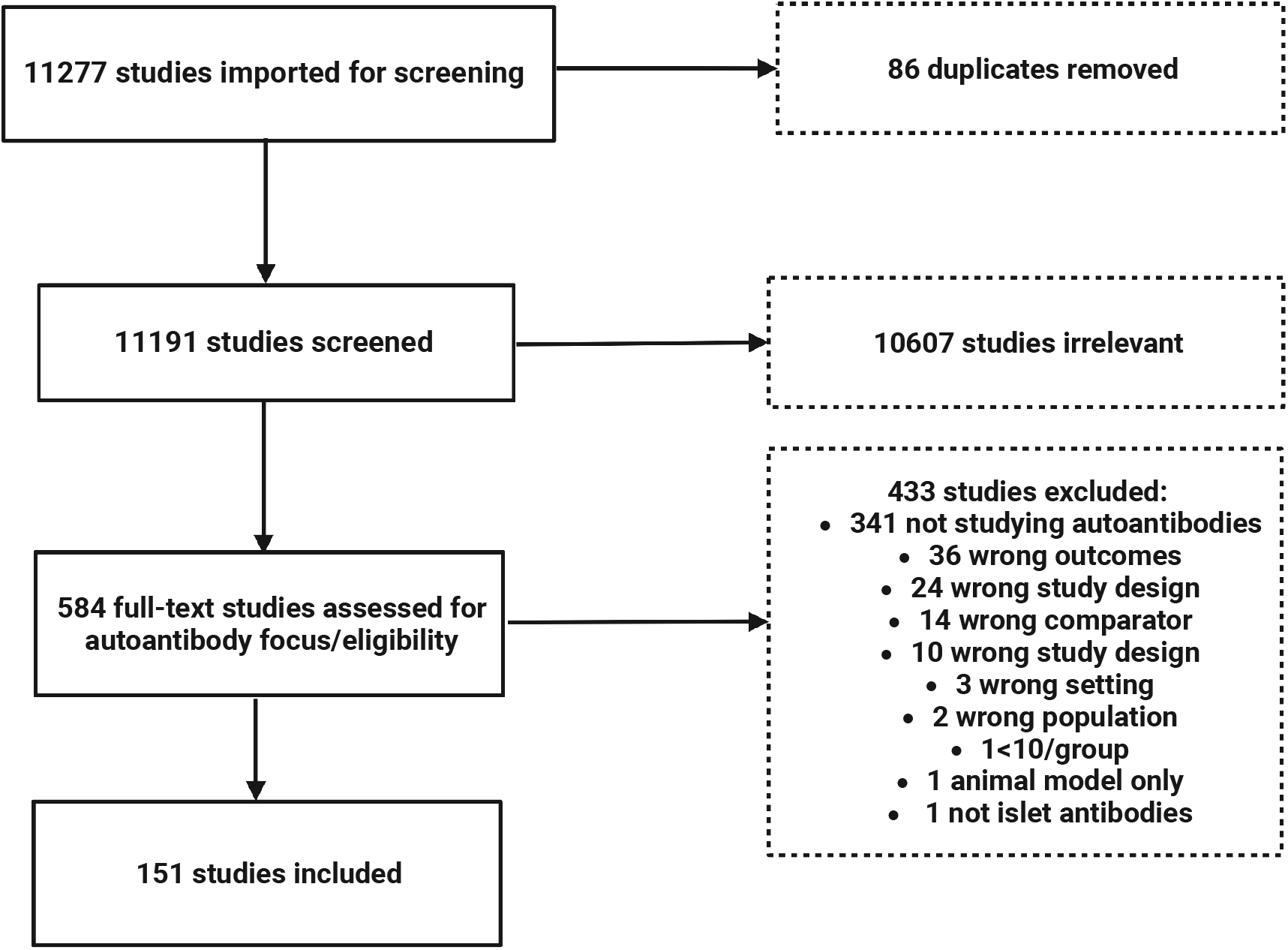
Consort diagram. Flow diagram for study selection.

All papers in the analysis assessed established islet autoantibodies (IAA, GAD, IA-2, ZnT8) or novel autoantibodies targeting other islet autoantigens. Islet autoantibodies were a primary focus for a majority of the papers identified (100/151, 66%), while others included autoantibody assessments as part of a larger precision analysis or clinical trial follow-up. Notably, less than half (65/151, 43%) reported using autoantibody assays that had been tested in a standardization program such as the islet autoantibody standardization program (IASP) (9). The most frequent autoantibody feature studied was autoantibody type/target protein (137/151, 91%), followed by autoantibody number (97/151, 64%), autoantibody titer (50/151,33%), age at seroconversion (39/151, 26%), rate of seroconversion from single to multiple autoantibodies (32/151, 21%), order of autoantibody appearance after seroconversion (28/151, 18%), novel islet autoantibody/epitope identification (13/151, 9%), and autoantibody affinity (6/151, 4%). Four of 151 papers (3%) assessed the use of different autoantibody assays to improve specificity of autoantibody testing.

Only 10/151 studies focused on a population that did not feature primarily European ancestry. In 110 studies, race and ethnicity were not reported, and those that did report race and ethnicity used inconsistent approaches to reporting (e.g., combined vs. separated race and ethnicity categories). Of studies that reported race and ethnicity, the median percentage of participants identifying as non-Hispanic white was 89% (IQR 84%-97%).

### Prior to diagnosis

The majority of the literature using autoantibody features to define heterogeneity in T1D focused on the period leading up to stage 3 T1D diagnosis (90/151, 60%). Studies included in the “prior to diagnosis” group are summarized in Supplemental Table 1. Median sample size was 510 (IQR 134-2239). Pediatric only populations were included in 61% (55/90), one study included only adults. The remainder (34/90, 38%) included pediatric and adult populations combined. Results were reported to have been impacted by age in 68% (61/90).

Assessment of islet autoantibody features during progression to T1D highlighted phenotypes characterized by age and genetic risk that were more clearly delineated with the addition of autoantibody type. Key recurring themes are summarized below.

#### Risk for progression to stage 3 T1D increases with autoantibody number

In 2013, a combined analysis of 3 large birth cohorts from different countries showed that T1D risk progressively increased with increasing numbers of positive autoantibodies. Of the 585 children who developed at least 2 autoantibodies, 84% developed type 1 diabetes within 15 years of follow-up (10). This appreciation that lifetime risk of diabetes progression nears 100% once multiple positive islet autoantibodies have developed informs the current T1D staging system (2). Since then, the impact of autoantibody number on risk of progression to stage 3 has been corroborated in numerous studies of additional cohorts (11–19).

#### Younger age at seroconversion results in faster progression to stage 3 T1D

Longitudinal assessment of over 2,500 children with genetic risk for T1D followed in the DAISY cohort revealed that speed of progression to T1D diagnosis is strongly correlated with age at seroconversion (20). These findings have been replicated in many subsequent T1D screening studies including an analysis of over 13,000 children from multiple birth cohorts (DAISY, DIPP, BABYDIAB, BABYDIET) in 2013 (10). The TrialNet Pathway to Prevention natural history study followed over 30,000 first and second degree relatives of individuals with T1D and revealed that frequency of seroconversion from single to multiple autoantibody positivity decreases with age (cumulative incidence 2% for age 10 and under, 0.7% for those over 10 years) (21). The clear relationship between younger age and faster progression was particularly strong prior to puberty (22). A recent analysis from TEDDY described an exponential decline in risk and rate of development of single and multiple autoantibodies with increasing age (23).

#### Islet autoantibody type (IAA, GAD, IA-2, ZnT8, ICA) influences progression

Autoantibody type can be used, in addition to autoantibody number, to stratify risk more precisely for T1D progression. Overall, IA-2 and ZnT8 positivity was associated with increased T1D pathogenicity. Multiple studies described an increased risk of progression from single to multiple autoantibody positivity or to stage 3 T1D associated with IA-2 positivity (14, 17, 18, 24–32). This was most clearly seen in pediatric populations, as IA-2 positivity was accompanied by development of other islet autoantibodies in 98% of the children followed in the BABYDIAB cohort (28).

However, when pediatric and adult populations were evaluated together, ZnT8 positivity was most commonly associated with development of other autoantibodies, and in single autoantibody positive subjects, if the single autoantibody was ZnT8, risk of progression to T1D was higher compared to single positivity for IAA, GAD, or IA-2 (33). Positivity for IAA and GAD was more often associated with decreased risk or slower progression to T1D. In analysis of pediatric and adult subjects, IAA or GAD positive first degree relatives progressed more slowly to T1D than double autoantibody positive subjects positive for IA-2 and ZnT8 (34). Reversion from single autoantibody positivity to autoantibody negativity was frequent for GAD and IAA, but not IA-2 and ZnT8 (13). Multivariate analysis of subjects < 20 years old showed that IA-2, IAA, ICA, and ZnT8 positivity, but not GAD, could all independently predict diabetes progression (33).

#### Order of autoantibody development varies by age and impacts risk for progression

Multiple longitudinal studies have shown that the first autoantibody to appear differs significantly depending on age of seroconversion. Analysis from the Finnish Type 1 Diabetes Prediction and Prevention (DIPP) study showed that in children 2 years old and younger, abnormal IAA titers most frequently develop first, while children ages 3-5 years more frequently seroconvert to GAD autoantibody positivity (35). A smaller analysis from the Diabetes Auto Immunity Study in the Young (DAISY) cohort found that higher IAA levels were associated with younger age at diagnosis, and that nearly all young children who progressed to T1D were IAA positive (36). Analysis of the BABYDIAB and BABYDIET pediatric cohorts also found that earliest autoantibody development (peak incidence 9 months) was most commonly development of a single IAA which progressed to multiple autoantibodies. Features linked to IAA as the first autoantibody include specific single nucleotide polymorphisms, male sex, father or siblings as a diabetic proband, introduction of probiotics at less than one month of age, and weight at 12 months (37). While less common, initial seroconversion to GAD resulted in slower progression to disease (compared to IAA, ICA, or IA-2) (12). Analysis of both pediatric and adult subjects combined revealed that the risk of progression from single to multiple autoantibodies decreased rapidly with increasing age when IAA was the first to develop. A decrease in risk with increasing age was also observed when GAD was the first to develop, but the risk reduction was less robust than that of IAA first (38).

#### The addition of autoantibodies improves performance of genetic risk stratification to predict progression

Highest genetic risk for T1D is associated with genes that encode MHC class II molecules (39). Multiple studies suggest that genetic risk stratification with other identified risk variants has the potential to be improved by the consideration of autoantibody features. While MHC class II-associated genetic risk is well defined, less is known about risk associated with MHC class I genes. In a study of Belgian adult and pediatric first-degree relatives who were carriers of the high-risk MHC class II HLA-DQ2/DQ8, risk was further increased by the presence of MHC class I HLA-A*24 if subjects were also positive for IA-2, but not if subjects were IA-2 negative. Additional screening for MHC class I HLA-B*18 with HLA-DQ2/DQ8 and HLA-A*24, and IA-2 and/or ZnT8 autoantibodies, increased the sensitivity of detecting rapid progressors (31). For single autoantibody positive relatives, combinations of autoantibody positivity and high-risk alleles improved risk predication, with younger age, HLA-DQ2/DQ8 genotype, and IAA positivity acting as independent predictors of more rapid seroconversion to multiple autoantibody positivity. The addition of autoantibody features to predict progression in genetically at-risk, multiple autoantibody positive relatives was less useful for this cohort, as most multiple autoantibody positive relatives progress to T1D within 20 years. Progression did occur more rapidly in the presence of IA-2 or ZnT8, regardless of age, HLA-DQ genotype, and autoantibody number (34). Among single and multiple autoantibody subjects, the non-HLA risk variant PTPN22 risk allele (T/T) was associated with faster progression to T1D after appearance of the first and second autoantibodies, indicating a higher risk subgroup, while the INS risk allele had no impact on the risk of progression to T1D (40). Genetic risk scores, calculated using multiple different genetic factors, were also shown to have the potential to be improved by the addition of autoantibody features. Redondo and colleagues using 30 T1D associated single-nucleotide polymorphisms, showed that positive predictive value of this score could be improved when the number of positive autoantibodies was also included in the model (41).

#### Positive predictive value of autoantibody titer and affinity varies by type

In addition to autoantibody type, autoantibody titers and affinities were also measured in many studies. However, only higher IAA and IA-2 titers and affinities have been shown to be linked to more rapid disease progression (42). In pediatric and adult first degree relatives followed in the DPT-1 cohort, IA-2 titers increase and GAD titers decrease in the years prior to T1D diagnosis (27). Similar findings were supported in young European children with HLA-DQB1-conferred disease susceptibility and advanced beta-cell autoimmunity where, in addition to young age, higher BMI SDS, and reduced first phase insulin response, higher IAA and IA-2 levels predicted T1D (43). In persistently autoantibody positive children in The Environmental Determinants of Diabetes in the Young (TEDDY) study, higher mean IAA and IA-2 levels, but not GAD levels were associated with increased T1D risk (11, 16). The addition of islet autoantibody features to existing metabolic measures alone will likely be less impactful in stratifying risk. In the DPT-1 study cohort, the addition of autoantibody titers did not improve a prediction model based on oral glucose tolerance testing, and IAA titers did not provide significant prediction value in subgroups with abnormal glucose tolerance (44).

#### Specific autoantibody assay methods impact risk stratification

Four papers assessed differences between traditional radiobinding (RBA) assays and newer electrochemiluminescent (ECL) assays. Overall, ECL assays had higher positive predictive value, were more sensitive, and defined seroconversion earlier than traditional RBA assays (45–47). This association was most pronounced in single autoantibody positive populations (46). In the DAISY cohort, only 3 of 11 single autoantibody positive children testing positive for ZnT8 by RBA were also positive for ZnT8 by ECL. All 3 progressed to T1D, suggesting that ECL assays may identify a subset of higher risk, single autoantibody positive individuals (47).

### At diagnosis

We identified 44 relevant studies that assessed the use of antibodies to define heterogeneous phenotypes at stage 3 T1D onset in the “at diagnosis” group (Supplemental Table 2). Median sample size was 561 (IQR 266-1036). Pediatric only populations were included in 28/44 (64%).

Multiple studies demonstrated differences in autoantibody type by age at diagnosis. Compared to children at onset, adults were less likely to be ICA positive or IA-2 positive; however, there were no differences in GAD positivity rates (48). In Chinese individuals with T1D, children with acute onset T1D showed higher prevalence of IA-2, ZnT8, and multiple autoantibody positivity than adults, and children diagnosed under 10 years had the highest frequency of IA-2 positivity and multiple antibody positivity (49). Children who develop autoantibodies and progress to T1D early in life have less functional beta-cell mass and higher rates of diabetic ketoacidosis (DKA) at diagnosis. No specific positive autoantibody type (GAD, IA-2, IAA, ZnT8) associated with DKA severity in children (50).

### After diagnosis

A total of 11 studies that assessed the use of autoantibodies to characterize progression after diagnosis were identified and summarized in Supplemental Table 3. All studies used C-peptide measures to assess endogenous insulin production. Median sample size was smaller for this group (247, IQR 129-367). The majority 9/11 (82%) included only pediatric populations.

Conclusions from this set of studies varied widely; however, a general theme was that less autoimmunity at diagnosis (as reflected by autoantibody titer and number) was more commonly associated with greater residual C-peptide. Persistent autoantibody negative status was associated with preserved residual C-peptide in multiple studies (51). Mean C-peptide 2 years post-diagnosis was correlated with absence of ICA or IAA at diagnosis in European ethnic groups (52). Higher antibody number at diagnosis was associated with lower rates of partial remission (53). In a study of new onset pediatric patients, when controlling for age, IAA positivity was associated with more rapid C-peptide decline post-diagnosis, while no relation was identified for GAD or IA-2 positivity (54).

### Treatment response

The “response to treatment” group included 13 primary randomized controlled trials of disease-modifying therapies tested prior to or at stage 3 T1D diagnosis (Supplemental Table 4). Three of these trials were designed to test agents targeting a specific antigen in a specific autoantibody positive group, with overall negative findings. The TrialNet oral insulin study, designed to test a subgroup identified as part of the DPT-1 study, where individuals with high IAA titers exhibited significant delay in time to diabetes compared to placebo, ultimately did not show an impact of oral insulin on time to diabetes in this population (55). Similarly, studies testing whether a GAD antigen-based immunotherapy was effective in GAD positive individuals did not see a treatment response (56, 57). Overall, responses to immunomodulatory therapies tended to differ by autoantibody type. Cyclosporin immune suppression tended to work more poorly in IA-2 positive individuals but reduced insulin requirements and increased C-peptide secretion in IA-2 negative individuals (58). In the teplizumab anti-CD3 prevention trial, treatment response was greater when ZnT8 was negative, and the presence or absence of other autoantibodies did not associate with clinical response (59). The B cell-depleting agent rituximab suppressed IAAs compared with placebo but had a much smaller effect on all other antibodies (60, 61); however, analysis of whether IAA positivity was associated with treatment response was not done. Importantly, all these trials tested combined pediatric and adult populations without considering age effects in autoantibody subgroup analyses, although none identified a positive impact of age itself on treatment efficacy.

### Risk of Bias Analyses

Reviewers performed assessments of specific metrics related to autoantibody assay quality as well as overall study design (Figure 2). Metrics to assess performance of autoantibody assays are shown in Figure 2A. Seventy-four percent (111/151) of studies applied the same assay to all participants tested; this either did not occur or was not clear in the remaining 26% (40/151) of studies. Methods used to measure autoantibodies were described in 58% (88/151) of papers. Of the 63 papers that did not give specific assay information typically either referenced another paper for methods (53/151, 35%) or did not focus on antibodies as a main outcome (8/151, 5%). About half (73/151, 48%) described characteristics of autoantibody assays utilized, such as sensitivity, specificity, and assay variation. Papers that did not most commonly referenced another paper for methods (43/151, 28%) or did not primarily focus on autoantibodies (20/151, 13%). Forty-four percent of total papers (65/151) referenced participation in an autoantibody standardization program. Of the 86 papers that did not mention this type of program, over half (54/86, 63%) primarily focused on autoantibodies.

**Figure 2.**
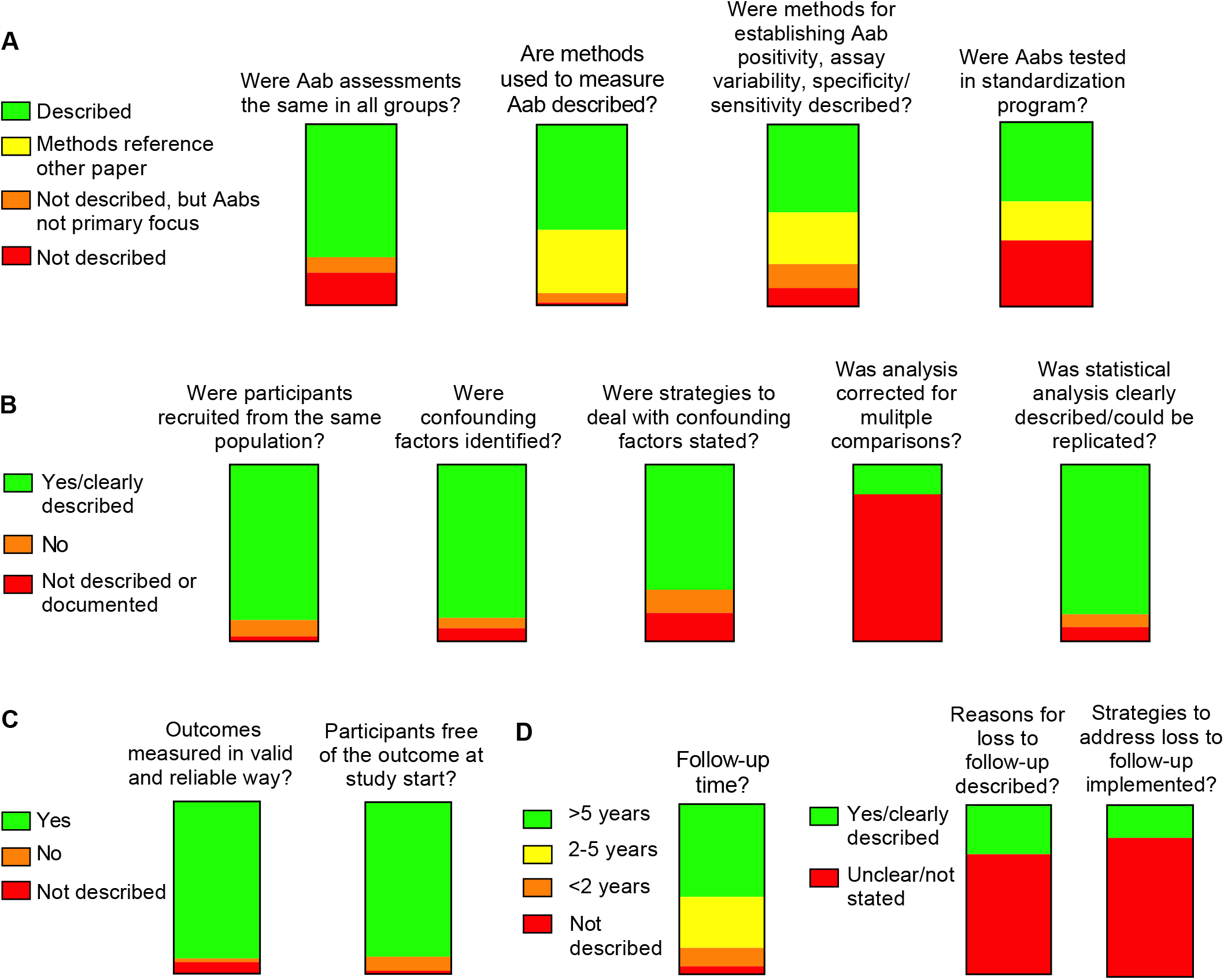
Quality assessment of literature search results. Graphical heat map of quality assessment questions surrounding A) autoantibody measurements, B) study design and analysis, C) outcome assessments, and D) study follow-up.

Quality assessments also touched on other aspects of study design. Reviewers judged that study participant groups were all recruited or identified from similar populations in most (133/151, 88%) papers. Confounding factors were presented in 87% (131/151) of papers, but only addressed in the analysis in 71% (107/151). Multiple analyses or comparisons were tested in the vast majority of papers (142/151, 94%), but only 24 of these (17%) described corrections for multiple comparisons. Statistical analyses were judged as clearly documented and able to be replicated in 85% (128/151) of papers, not clearly documented in 8% (12/151) and documented but with concerns raised for approach in 7% (11/151).

Diagnosis of T1D was measured as an outcome in 91% of papers and of these, 125 (91%) included valid and reliable measures of T1D diagnosis. For the 110 papers with dichotomous outcomes over a period of follow-up, 90% (99/110) clearly described methods to ensure that participants were free of the outcome at study start. 110 studies included longitudinal follow-up. Specific descriptions of the follow-up period were typically included; this was most commonly over > 5 years (60/110, 55%), with 30% (33/110) followed for 2-5 years, and 11% (12/110) followed for < 2 years. Duration of follow-up was clearly described in 105/110 (95%). Loss to follow up was much less commonly described. Specifics were only included in 29% (32/110) of applicable papers, and strategies to address loss to follow-up were only described in 19% (21/110) of applicable papers.

## Discussion

We hypothesized that islet autoantibodies could be used to identify and define specific phenotypes prior to, at, and after stage 3 T1D diagnosis, and in response to disease modifying therapy. We systematically reviewed the application of antibody measurements to define heterogeneity at diagnostic timepoints before and after the onset of clinically symptomatic disease. We reviewed 151 papers published over the past 10 years meeting inclusion criteria and identified recurring themes in the literature. Strikingly, nearly all studies identified assessed antibody features prior to diagnosis, suggesting that overall, the application of antibody features to precision medicine diagnostics will be most impactful on defining differences in T1D phenotypes during this period of disease development.

Although multiple individual features (immune signatures, genetics, metabolic measures) could be applied to differentiate disease phenotypes, in this effort, we chose to focus on autoantibodies because standardized measures are currently available and their implementation as precision diagnostic tools in T1D has the potential to be rapidly implemented. As a well-established marker of islet autoimmunity, autoantibodies benefit from prior harmonization efforts, existing standardization workshops that compare assays using clinical samples, and for the most-established assays, easy accessibility to clinicians (5–8). Indeed, the application of autoantibody number as a precision diagnostic tool that stratifies future disease risk has gained traction within the T1D research field with the T1D staging system and has already been used in research to define stages of T1D (2). Our review supports broad application of T1D staging using autoantibody number to guide individuals and clinicians on T1D risk. Furthermore, our findings support conclusions that have been drawn by others (62) that additional antibody features could be utilized to more precisely stratify the current staging paradigm.

Our analysis supports existing evidence of the strong impact of age on heterogeneity of T1D development. Specifically, our analysis confirms that younger age at seroconversion increases risk and rate for progression to stage 3 T1D, and this age-related risk can be further stratified using islet autoantibody type and titer. The significant impact of age when considering use of autoantibodies to stratify risk suggests that a) recommendations for use of autoantibodies in screening and prediction studies will need to consider stratification by age groups, and b) that the analytic approach to autoantibody studies should include adjustments for age.

While most studies on autoantibodies in the prediabetes period focused on autoantibody number, type, or timing of seroconversion, fewer studies that passed our criteria for review assessed the immune responses that drive these changes. Therefore, there is continued need to understand how and when tolerance is broken in the context of clinical trials and how this leads to heterogeneous phenotypes. The few studies that did assess immune signatures in multiple autoantibody positive relatives revealed both proinflammatory and partially regulated (protective) phenotypes, which were also associated with autoantibody number. Interestingly, autoantibody negative relatives were characterized by the partially regulated phenotype (63), suggesting that progression to T1D may be the result of insufficient suppressive mechanisms, rather than differences in antigen targets. Immunoregulatory signatures were also identified in high risk siblings of subjects with T1D who were autoantibody negative (64), though this study was excluded from our review due to sample size.

Precision diagnostics has particular utility in stratifying risk beyond autoantibody number in single autoantibody positive individuals, a group that is considered lower risk overall, and consequently, often do not meet inclusion criteria for clinical trials that require multiple positive autoantibodies. Studies of ECL vs RBA assays suggest that ECL assays can identify a subset of higher risk, single autoantibody positive individuals. Autoantibody type was identified in this review as a common approach to stratify risk among single autoantibody positive individuals. For example, given the rarity of IA-2 as the initial autoantibody at seroconversion in birth cohorts, individuals who are cross-sectionally single autoantibody positive for IA-2 may reflect a higher risk group that has reverted to single autoantibody positivity, and are at higher risk of progression to multiple autoantibody positivity and ultimately T1D.

Evidence for use of antibodies at, after, and in response to disease-modifying therapies was less robust, and far fewer studies were identified. At diagnosis, the presence or absence of specific islet autoantibodies was also correlated with age, which might be expected given the differences in autoantibody presentation at seroconversion. However, multiple studies suggested that the primary autoantibodies at seroconversion had often disappeared at the time of diagnosis; this is an open question in the field. While some evidence suggested that declining islet autoantibody titers and numbers after diagnosis corresponded to preserved residual C-peptide, we did not find convincing evidence to support the use of islet autoantibodies to define heterogeneity in metabolic outcomes after stage 3 diagnosis. Evidence for use of islet autoantibody features to predict responses to disease modifying therapies was modest. We postulate that this is likely secondary to the epitope spreading and neoantigen expansion that accompanies T1D disease progression, making the impact of a specific antigen (and its corresponding autoantibody) less significant by the time an individual has reached more advanced stages of disease.

Of note, our initial search strategy that targeted papers with combined use of MeSH (Medical Subject Headings) terms for “Precision Medicine” and “Type 1 Diabetes” identified only a small number of papers which were predominantly commentaries or reviews with very few original research articles. This likely represents the relatively recent application of precision medicine concepts to the field as well as a broader issue surrounding nebulous definitions of precision medicine (65). Moving forward, inclusion of “Precision Medicine” as a MeSH term in papers focused on T1D heterogeneity, stratification, or endotypes will be critical to allow researchers to easily access relevant studies in this area.

Many of the studies we reviewed emanated from prospective longitudinal cohort studies either from prevention trials or natural history cohorts, such as DPT-1, DAISY, BABYDIAB, TEDDY, Fr1da, DIPP, and the TrialNet Pathway to Prevention study. Likely related to this, overall, outcomes were judged to be reliably ascertained, and participants had substantial durations of follow-up (55% documented follow-up beyond 5 years). These qualities highlight the exceptional value that natural history cohorts have brought to the field of T1D precision medicine overall.

However, our review has also highlighted quality concerns that will benefit from being addressed moving forward. An area that is particularly high yield is the use of autoantibody standardization workshops aimed at improving the performance and concordance of immunoassays used to measure islet autoantibodies. Despite standardization programs being available throughout the timeframe studied in this review, participation in theses workshops was not uniformly referenced, even among papers with autoantibodies as a primary focus. Especially with more novel assays, clear reporting of methods and validation efforts are critical to the reproducibility of findings (5). The fact that the framework has already been set to do this through the establishment of existing standardization programs makes improved and more consistent participation in these workshops “low hanging fruit” for improvement of study quality.

Loss to follow-up in longitudinal studies was not frequently documented. While analysis strategies frequently addressed differences in follow-up duration, systematic differences in loss to follow-up amongst different populations could theoretically still impact findings. Additionally, given the frequent reporting of interactions of autoantibody findings with age, consideration of relationships with age and other confounding factors is critical. This did not appear to be addressed in 30% of papers reviewed.

The vast majority of studies assessed emanated from cohorts that were composed of groups of individuals of primarily European ancestry. Specific reporting on race and ethnicity were uncommon (only present in about 1/4 of papers) and were inconsistently applied. Validation of antibodies as a tool for precision diagnosis across diverse populations, such as has been performed with genetic T1D risk scores (66) will be important to ensure broader applicability.

There are some limitations to this analysis. Mainly for review feasibility, we limited our search to a 10-year period and to outcomes related to islet autoantibodies. Because of this, important papers in the field that did not meet inclusion criteria were not included as part of this review. For example, the Fr1da-study group recently published data that showed IA-2 positivity and titer, in combination with hemoglobin A1c and OGTT glucose values, could be used to generate a progression likelihood score that effectively identified presymptomatic multiple autoantibody positive children at very high risk of progression to clinical disease (67). However, this study is not included because it was published several months after the date of the literature search. Given differences in exposures, outcomes, and study conditions, we were not able to perform a meta-analysis on this topic, but instead chose to systematically review the state of the literature.

Notwithstanding these limitations, overall, our findings suggest that islet autoantibodies are likely to be most useful to define T1D heterogeneity prior to clinical diagnosis, supporting prior efforts to use autoantibodies as part of precision T1D staging. Further benefit may be gained by their incorporation into risk scores that include features beyond autoantibody number and also consider age and genetics. Moving forward, thoughtfully designed, prospective analyses to test these relationships with disease-modifying therapies would help to further apply these observations and develop precision medicine approaches to T1D prevention. Additionally, systematic review of other individual features, such as genetics, metabolic function, and other immune findings, will likely provide further insight into the current evidence for strategies to apply these features to precision T1D diagnostics.

## Data Availability

All data produced in the present study are available upon reasonable request to the authors.

