## Supplemental material for "Precision Diagnostics: Using Islet Autoantibodies to Characterize Heterogeneity in Type 1 Diabetes"

### Supplemental Figure 1. Search strategy.

**2022-06-14**

**Pubmed**

**#1**
**("Precision Medicine"[Mesh] OR Subtype*[Title/Abstract] OR heterogeneity[Title/Abstract] OR heterogeneity[Title/Abstract] OR endotype*[Title/Abstract] OR personalized[Title/Abstract] OR tailored[Title/Abstract] OR strat*[Title/Abstract] OR subgroup*[Title/Abstract] OR variability[Title/Abstract] OR phenotype*[Title/Abstract] OR pattern*[Title/Abstract] OR predict*[Title/Abstract] OR stage [Title/Abstract] OR variant*[Title/Abstract] OR predict*[Title/Abstract] OR risk*[Title/Abstract] OR genetic risk score*[Title/Abstract] OR signature*[Title/Abstract] OR ("genetic predisposition to disease"[MeSH Terms]))**
**=8216498**

**#2**
**"Diabetes Mellitus, Type 1 "[Mesh]**
**=83221**

**#3**
**#1 AND #2**
**=29779**

**#4**
**(review*[Title/Abstract]) OR (review*[Publication Type])**
**=4132084**

**#5**
**#3 NOT #4**
**=23830**

**#6**
**#7 Filters: Humans, English, from 2011 – 2022**
**=9941**

Supplemental Table 1. Autoantibody features characterize progression before T1D diagnosis.

| Study | n | Age group | Population Studied* | Autoantibody Feature Assessed | Age impact? | Findings |
|---|---|---|---|---|---|---|
| Eising 2011[1] | 4,042 | Pediatric and adult | • Other: individually matched case-controls for all Danish T1D patients born between 1981 and 2002 and dx with T1D before May 1, 2004 | • Aab number<br>• Aab titer<br>• Aab type<br>• Age at Aab seroconversion | Not available | • Infants who were GAD and IA-2+ at birth had increased risk (HR 4.55) for T1D after controlling for parental diabetes and HLA-DQB1 alleles.<br>• HR for T1D diagnosis increases with titers of GAD and IA-2 at birth. |
| Sosenko 2011 (DPT-1)[2] | 99 | Pediatric and adult | • FDR<br>• Second degree relative<br>• Other: No relative but ICA+ and AGT | • Aab number<br>• Aab titer<br>• Aab type | Not available | • IA-2 titers increase in the years prior to T1D diagnosis.<br>• GAD titers decrease in the years prior to T1D diagnosis. |
| Steck 2011 (DAISY)[3] | 2542 | Pediatric | • High genetic risk<br>• FDR<br>• Second degree relative<br>• Single Aab+<br>• Multiple Aab+<br>• Other: undefined relative | • Aab number<br>• Aab titer<br>• Aab type<br>• Aab timing | Yes | • In children with high-risk genotypes, 89% who progressed to T1D had multiple Aabs.<br>• Age at T1D diagnosis is strongly correlated with age at seroconversion and levels of IAA. |
| Vehik 2011 (TrialNet)[4] | 32,845 | Pediatric | • FDR<br>• Second degree relative | • Aab number<br>• Aab type<br>• Aab timing<br>• Age at Aab seroconversion<br>• Order of Aab seroconversion | Yes | • Of >30,000 FDR who were initially Aab-, 205 seroconverted to GAD, 155 to IAA, and 53 to ICA<br>• Risk of IAA and GAD seroconversion decreased with increasing age.<br>• Cumulative Aab seroconversion was 2% for age 10 and under, 0.7% for those over 10 years. |
| Beyan 2012[5] | 115 | Pediatric and adult | • Single Aab+<br>• Other: ICA+ from general population | • Aab number<br>• Aab type | Not available | • In a cohort of 115 ICA+ schoolchildren, only ZnT8 contributed to prediction of diabetes.<br>• In cox regression model, ZnT8 independently predicted diabetes. |

Supplemental Table 1. Autoantibody features characterize progression before T1D diagnosis.

| | | | | | | |
|---|---|---|---|---|---|---|
| Krause 2012 (BABYDIAB)[6] | 50 | Pediatric | • FDR<br>• Single Aab+<br>• Multiple Aab+ | • Aab affinity | Yes | • Low affinity IA-2 Aabs are rare.<br>• IA-2 affinity is high from first detection of Aabs.<br>• IA-2 affinity did not stratify T1D risk.<br>• IA-2 preceded or was accompanied by other islet Aabs in 98% of at risk children. |
| Lempainen 2012 (DIPP)[7] | 249 | Pediatric | • Single Aab+<br>• Multiple Aab+ | • Aab number<br>• Aab type | Not available | • PTPN22 risk allele (T/T) but not INS risk allele was associated with faster progression after appearance of first and second Aabs. |
| Long 2012 (ENDIT)[8] | 526 | Pediatric and adult | • FDR<br>• Single Aab+<br>• Multiple Aab+<br>• Other: general population ICA+ | • Aab type | Yes | • In Aab+ relatives, ZnT8 improved prediction in relatives at low genetic risk of T1D and > age 20 yrs.<br>• In individuals with ICA+ alone, IA-2 negative relatives, additional Aabs, younger relatives, and relatives with high or intermediate genetic risk, ZnT8 did not improve risk prediction. |
| Parikka 2012 (DIPP)[9] | 1320 | Pediatric | • High genetic risk<br>• Single Aab+<br>• Multiple Aab+ | • Aab number<br>• Aab titer<br>• Aab type<br>• Aab timing<br>• Age at Aab seroconversion<br>• Order of Aab seroconversion | Yes | • Early initiation of autoimmunity and rapid increases in Aab titers strongly predict progression to T1D before puberty.<br>• Incidence of seroconversion peaked at 1 year<br>• AAb titers were higher 3 to 6 month after seroconversion in progressors to T1D vs nonprogressors. |
| Xu 2012 (DPT-1)[10] | 339 | Pediatric and adult | • FDR<br>• Second degree relative<br>• Other: ICA+ and abnormal glucose tolerance/abnormal FPIR | • Aab titer<br>• Aab type | No | • Adding Aab titer markers to OGTT prediction model for T1D did not improve accuracy.<br>• IAA titer did not provide significant prediction value in subgroups with AGT and had poor predictive performance. |
| Yu 2012 (TrialNet)[11] | 1770 | Pediatric and adult | • High genetic risk<br>• FDR<br>• Second degree relative<br>• Single Aab+<br>• Multiple Aab+<br>• Other: Third degree relative | • Aab number<br>• Aab type | Yes | • ZnT8 positivity is associated with presence of other Aabs.<br>• In single Aab+ relatives, risk for T1D progression is higher if Aab is ZnT8 compared to other Aabs.<br>• Multivariate analysis showed age <20 years, IA-2A, IAA, ICA, and ZnT8 independently predicted diabetes, GAD did not. |

Supplemental Table 1. Autoantibody features characterize progression before T1D diagnosis.

| | | | | | | |
|---|---|---|---|---|---|---|
| Achenbach 2013 (BABYDIAB)[12] | 47 | Pediatric | • FDR<br>• Multiple Aab+<br>• Other: infants born to mother or father with T1D | • Aab number<br>• Aab type<br>• Aab timing | Yes | • In multiple Aab+ children, IA-2 development was delayed in slow vs. rapid progressors. |
| Andersson 2013 (DiAPREV-IT)[13] | 47 | Pediatric | • High genetic risk<br>• Multiple Aab+ | • Aab number<br>• Aab titer<br>• Aab type | No | • With the exception of ZnT8, Aab number and titer were not associated with glucose metabolism in multiple Aab positive individuals.<br>• In children with high genetic risk and multiple autoantibodies, impaired glucose metabolism was associated with a higher frequency and higher titer of ZnT8. |
| Gorus 2013 (BDR)[14] | 394 | Pediatric and adult | • FDR | • Aab number<br>• Aab type | Yes | • Rates of progression were independent of age in IA-2+ and/or ZnT8+ FDR.<br>• Rates of progression were decreased with age if only GAD and/or IAA were present.<br>• In 10-39yo FDR, screening for IA-2 and ZnT8 alone identified 78% rapid progressors, (vs 75% if positive for at least 2 antibodies among IAA, GAD, IA-2 and ZnT8 and 62% without testing for ZnT8). |
| Ilonen 2013 (DIPP)[15] | 520 | Pediatric | • High genetic risk | • Aab type<br>• Age at Aab seroconversion<br>• Aab order | Yes | • Aab type that appears first differs significantly depending on age.<br>• IAA first peaks at age 2.<br>• GAD first peaks at age 3-5.<br>• IA-2 rarely appears first. |
| Larsson 2013 (TrialNet, DiPiS)[16] | 395 | Pediatric and adult | • High genetic risk<br>• FDR | • Novel Aab/epitope | Yes | • FDR who progressed to T1D had lower anti-idiotype to GAD compared to those who did not progress. |

Supplemental Table 1. Autoantibody features characterize progression before T1D diagnosis.

| | | | | | | |
|---|---|---|---|---|---|---|
| Mbunwe 2013 (BDR)[17] | 288 | Pediatric and adult | • FDR<br>• Single Aab+<br>• Multiple Aab+ | • Aab number<br>• Aab type | Yes | • Compared with nonprogressors, rapid progressors (within 5 years of first Aab+) were younger and more frequently IA-2+, ZnT8+, multiple Ab+, and/or HLA-DQ2/DQ8+ .<br>• HLA-A*24, HLA-DQ2/DQ8, and positivity for IA-2 and/or ZnT8 were associated with development of T1D .<br>• HLA-A*24 increased progression in presence of HLA-DQ2/DQ8 or IA-2+ with or without ZnT8, but not in their absence.<br>• T1D risk and diagnostic sensitivity were higher in IA-2+ +/- ZnT8+ relatives than when HLA-DQ2/ DQ8 or HLA-A*24 carrier status alone was examined. |
| Mbunwe 2013 (BDR)[18] | 288 | Pediatric and adult | • FDR<br>• Single Aab+<br>• Multiple Aab+ | • Aab type | "Borderline significance" | • In Aab+ FDR, IA-2+ and/or ZnT8+ predicted diabetes development within 5 years.<br>• Additional screening for HLA-B*18 in addition to HLA-DQ2/DQ8 and HLA-A*24, and presence of IA-2 and/or ZnT8 Aabs significantly increases the sensitivity of detecting rapid progressors. |
| Siljander 2013 (DIPP)[19] | 218 | Pediatric | • High genetic risk<br>• Multiple Aab+ | • Aab number<br>• Aab titer<br>• Aab type<br>• Aab timing<br>• Age at Aab seroconversion | Yes | • Young age, higher BMI SDS, reduced FPIR and higher levels of IAA and IA-2 predicted T1D in young children with HLA-DQB1-conferred disease susceptibility and advanced beta-cell autoimmunity. |
| Sosenko 2013 (TrialNet)[20] | 784 | Pediatric and adult | • FDR<br>• Second degree relative<br>• Single Aab+<br>• Multiple Aab+ | • Aab number<br>• Aab titer<br>• Aab type | Yes | • Strong association of the progression to T1D with the composite of the levels from the five autoantibodies before and after an adjustment for their positivity or negativity.<br>• The autoantibody risk score was strongly predictive of T1D.<br>• The combination of the autoantibody risk score and the previously validated Diabetes Prevention DPTRS predicted T1D more accurately than either alone. |

Supplemental Table 1. Autoantibody features characterize progression before T1D diagnosis.

| | | | | | | |
|---|---|---|---|---|---|---|
| Yu 2013 (DAISY)[21] | 47 | Pediatric | • High genetic risk<br>• Single Aab+<br>• Multiple Aab+ | • Aab type<br>• Age at Aab seroconversion<br>• Order of Aab seroconversion<br>• Novel Aab/epitope | Yes | • Younger age at seroconversion corresponds to younger age at diagnosis.<br>• Higher IAA levels associated with younger age at diagnosis.<br>• Nearly all young children progressing to diabetes are IAA positive.<br>• ECL-IAA assay is more sensitive that defines seroconversion earlier than RBA-IAA assay. |
| Ziegler 2013 (DAISY, DIPP, BABYDIAB, BABYDIET)[22] | 13,377 | Pediatric | • High genetic risk<br>• FDR | • Aab number<br>• Aab type<br>• Aab timing<br>• Age at Aab seroconversion | Yes | • Risk of progression to T1D after 10 years was 70% for multiple Aab+,14% for single Aab+ children.<br>• Risk of progression to T1D by 15 years was 0.4% in Aab- children.<br>• Progression to T1D in children with multiple Aab is faster for children with seroconversion < 3 years. |
| Arif 2014[23] | 105 | Pediatric | • FDR<br>• Single Aab+<br>• Multiple Aab+<br>• New onset | • Aab number<br>• Aab type | Yes | • Two distinct immune phenotypes were identified among individuals with new onset T1D and high risk siblings with multiple Aab: half with proinflammatory (IFN-gamma+, multiple Aab+) and half with partially regulated (IL-10+, pauci-aab+) phenotypes.<br>• In Aab- siblings with low T1D risk, IL-10-mediated autoreactivity is frequently detected. |
| Bender 2014 (Karlsburg Type 1 Diabetes Risk Study)[24] | 97 | Pediatric | • Single Aab+<br>• Multiple Aab+ | • Aab affinity | Not available | • GAD affinity was high in most multiple Aab positive children.<br>• There was a wide range of affinity in single GAD positive children, but those who developed multiple antibodies on follow-up had high affinity GAD.<br>• 88% children who developed T1D had high-affinity GAD prior to onset. |
| Elding Larsson 2015 (DiAPREV-IT)[25] | 50 | Pediatric | • FDR<br>• Multiple Aab+<br>• Other: general population | • Aab number<br>• Aab titer<br>• Aab type<br>• Aab timing | No | • IA-2 levels above median increased the risk for progression to T1D. |

Supplemental Table 1. Autoantibody features characterize progression before T1D diagnosis.

| | | | | | | |
|---|---|---|---|---|---|---|
| Giannopoulou 2015 (BABYDIAB, BABYDIET)[26] | 2,441 | Pediatric | • FDR | • Aab number<br>• Aab type<br>• Aab timing<br>• Age at Aab seroconversion<br>• Order Aab seroconversion | Yes | • In children with FDR/parents with T1D, seroconversion with single IAA was more common than multiple Aab+.<br>• Multiple Aab+ in children with FDR/parent with T1D was more common than single GAD+.<br>• Earliest Aab development was seen in children with single IAA that progressed to multiple Aabs or with persistent high-affinity single IAA (peak incidence 9 months).<br>• For children who underwent multiple Aab seroconversion, peak incidence was 2 years.<br>• For children who first seroconverted to GAD and later other Aabs, peak incidence was 5 years.<br>• Seroconversion to low-affinity IAA or persistent single GAD occurred at a low incidence after the age of 9 months.<br>• Progression to T1D occurred in >50% of children within 10 years in all groups that developed multiple islet autoantibodies and in 44% of children with persistent single high-affinity IAA or persistent single GAD antibody.<br>• There was a higher incidence of early insulin autoimmunity in children with HLADR4-DQ8/DR4-DQ8 and DR3/DR4-DQ8 genotypes. |
| Lempainen 2015 (DIPP)[27] | 521 | Pediatric | • High genetic risk<br>• FDR | • Aab number<br>• Aab type<br>• Order of Aab seroconversion | Yes | • IKZF4-ERBB3 associated with GAD as first Aab, INS genotype associated with IAA as first Aab. |
| Steck 2015 (TEDDY)[28] | 577 | Pediatric | • High genetic risk | • Aab number<br>• Aab titer<br>• Aab type<br>• Aab timing<br>• Age at Aab seroconversion | Yes | • Cumulative incidence of diabetes by 5 years since the appearance of the first Aab differed significantly by the number of positive Aabs.<br>• Higher mean IAA and IA-2 levels were associated with an increased risk T1D in children who were persistently Aab+.<br>• Mean GAD level did not significantly affect the risk of diabetes. |

Supplemental Table 1. Autoantibody features characterize progression before T1D diagnosis.

| | | | | | | |
|---|---|---|---|---|---|---|
| Williams 2015 (BOX)[29] | 283 | Pediatric and adult | • High genetic risk | • Aab type<br>• Novel Aab/epitope | Not available | • In FDRs positive for GAD(1-585), those also positive for truncated GAD(96-585) had higher rates of progression to T1D. |
| Bingley 2016 (TrialNet)[30] | 983 | Pediatric and adult | • FDR<br>• Second degree relative<br>• Single Aab+ | • Aab number<br>• Aab type<br>• Aab timing<br>• Age at Aab seroconversion<br>• Order of Aab seroconversion | Yes | • Younger age (<13yrs) male sex, white race, and higher risk HLA class II genotype associated with higher risk of multiple Aab development.<br>• Aab type (GAD, IA-2, IAA) was not associated with differences in risk of multiple Aab development.<br>• Relatives who progressed from single to multiple Aab+ had higher risk of T1D than those who initially screened as multiple Aab+. |
| Endesfelder 2016 (BABYDIAB)[31] | 88 | Pediatric | • FDR | • Aab number<br>• Aab titer<br>• Aab type<br>• Aab timing<br>• Order of Aab seroconversion | No | • FDR with only 2 Aabs (and did not develop more) progressed more slowly.<br>• FDR with more than 2 Aabs but lacking or losing IAA over time progressed more slowly. |
| Fouts 2016 (TrialNet)[32] | 1,287 | Pediatric and adult | • FDR<br>• Second degree relative<br>• Single Aab+<br>• Multiple Aab+ | • Comparison of Aab assays | Not available | • Subjects ECL positive for GAD and IAA had a risk of progression to diabetes within 6 years of 58% compared with 5% for the ECL-negative subjects.<br>• Adding ECL to the DPTRS improved the ROC curves with AUC of 0.83 ($p<0.0001$).<br>• ECL assays improved the ability to predict time to diabetes in Aab+ relatives. |
| Steck 2016 (DAISY)[33] | 91 | Pediatric | • High genetic risk<br>• FDR | • Aab number<br>• Aab titer<br>• Aab type<br>• Age at Aab seroconversion<br>• Order of Aab seroconversion | Yes | • Progression to stage 3 T1D in high genetic risk FDR was more common if first Aab appeared at younger age.<br>• Progression to stage 3 T1D in high genetic risk FDR was more common if IAA titers were high. |
| Steck 2016 (TrialNet)[34] | 100 | Pediatric and adult | • FDR<br>• Second degree relative<br>• Single Aab+ | • Aab affinity<br>• Aab type<br>• Comparison of Aab assays | Yes | • Among either single GAD or single IAA subjects, those who were positive in the ECL assay showed higher affinity at the initial visit, and affinity results stayed consistent over time.<br>• No converting events from low to high or high to low affinity were seen over time. |

Supplemental Table 1. Autoantibody features characterize progression before T1D diagnosis.

| | | | | | | |
|---|---|---|---|---|---|---|
| Vehik 2016 (TEDDY)[35] | 596 | Pediatric | • High genetic risk<br>• Single Aab+<br>• Multiple Aab+ | • Aab number<br>• Aab titer<br>• Aab type<br>• Age at Aab seroconversion<br>• Order of aab seroconversion<br>• Other: Aab reversion | Yes | • Reversion for GAD and IAA was frequent, but restricted to single Aab+ children, rare in multiple Aab+ children.<br>• Most reversion of single Aab+ children occurred within 2 years of seroconversion.<br>• Reversion was associated with HLA, age, and decreasing Aab titer.<br>• T1D risk remained high in multiple Aab+ children, even if a single Aab reverted. |
| Xu 2016 (TrialNet)[36] | 3270 | Pediatric and adult | • High genetic risk<br>• FDR<br>• Second degree relative<br>• Single Aab+<br>• Multiple Aab+<br>• Other: dysglycemia | • Aab number<br>• Aab titer<br>• Aab type<br>• Aab timing<br>• Age at Aab seroconversion | Yes | • Age and GAD titers defined three risk classes for progression from single to multiple Aab: 5 year risk for T1D was lowest (11%) in subjects >16 yrs with low GAD titers, higher in subjects <16 yrs with low GAD titers (29%), and highest (45%) for subjects with high GAD titers, independent of age.<br>• Progression to dysglycemia associated with IA-2 titers, 2h glucose, and fasting C-peptide.<br>• Progression to T1D associated with Aab number, peak C-peptide, HbA1c, and age. |
| Bosi 2017 (TrialNet)[37] | 994 | Pediatric and adult | • FDR<br>• Second degree relative<br>• Single Aab+ | • Aab number<br>• Aab type | Yes | • There is a rapid decrease in risk of progression from single to multiple Aabs with increasing age if IAA is primary Aab.<br>• There is a slower decrease in risk of progression from single to multiple Aabs with increasing age if GAD is primary Aab. |
| Frohnert 2017 (DAISY)[38] | 207 | Pediatric | • High genetic risk<br>• FDR<br>• Single Aab+<br>• Multiple Aab+ | • Aab number<br>• Aab type<br>• Aab timing<br>• Age at Aab seroconversion<br>• Order of Aab seroconversion | Yes | • Late onset of islet autoimmunity (age >8 years) was more common in African American and Hispanic individuals, more likely to present with a single Aab, was associated with increased rate of reversion to Aab- and slower progression to T1D.<br>• About half of those with late-onset islet autoimmunity progress to multiple Aab+ and T1D in adolescence/early adulthood.<br>• Aab profiles differed by age of onset (IAA being more prominent in the early-onset group, GAD more prominent in the late-onset group). |

Supplemental Table 1. Autoantibody features characterize progression before T1D diagnosis.

| | | | | | | |
|---|---|---|---|---|---|---|
| | | | | | | • DR3/4 HLA type was associated with a median time to reversion that was over three times longer than that for other HLA types. |
| Gorus 2017 (BDR)[39] | 462 | Pediatric and adult | • High genetic risk<br>• FDR<br>• Single Aab+<br>• Multiple Aab+ | • Aab number<br>• Aab titer<br>• Aab type<br>• Aab timing<br>• Age at Aab seroconversion<br>• Order of Aab seroconversion<br>• Novel Aab/epitope | Yes | • Multiple Aab+ progression more rapidly than single Aab+.<br>• IAA and GAD+ relatives progressed more slowly than double Aab+ individuals with IA-2 and/or ZnT8.<br>• In multiple Aab+ relatives, only IA-2 or ZnT8 were only independent predictors of more rapid progression.<br>• In single Aab+ relatives, younger age, HLA-DQ2/DQ8genotype, and IAA were independent predictors of seroconversion to multiple Aab+.<br>• In single Aab+ relatives, the time to multiple Aab+ increases with age and absence of IAA and HLA-DQ2/DQ8 genotype.<br>• Majority of multiple Aab+ relatives progress to T1D within 20 years; this occurs more rapidly in the presence of IA-2 or ZnT8, regardless of age, HLA-DQ genotype, and Aab number. |
| Krischer 2017 (TEDDY)[40] | 412 | Pediatric | • High genetic risk<br>• Multiple Aab+ | • Aab number<br>• Aab type<br>• Age at Aab seroconversion | Yes | • Age at developing multiple Aabs as first-appearing indication of seroconversion were associated with more rapid progression from multiple Aab+ to T1D. |

Supplemental Table 1. Autoantibody features characterize progression before T1D diagnosis.

| | | | | | | |
|---|---|---|---|---|---|---|
| Köhler 2017 (TEDDY)[41] | 613 | Pediatric | • High genetic risk<br>• FDR<br>• Single Aab+<br>• Multiple Aab+ | • Aab number<br>• Aab titer<br>• Aab type<br>• Aab timing<br>• Age at Aab seroconversion<br>• Order of Aab seroconversion | | • In children with high genetic risk, IAA, GAD, or both, were present at seroconversion, IA-2 occurred later.<br>• IAA seroconversion occurred at lower median age.<br>• IAA titers declined after an initial increase; GAD and IA-2 titers increased after seroconversion and remained stable.<br>• Risk of progression to T1D increased with titer for all Aabs; this association was time constant for IA-2, but decreased over time for IAA and GAD.<br>• Titers over time were lower for subjects who seroconverted at an older age and higher if the Aab appeared at initial seroconversion, and if other Aabs were present.<br>• For each Aab, a higher age at seroconversion was associated with lower risk of progression to T1D.<br>• A higher association was observed between Aab trajectories and time to T1D for IAA+ subjects and the DR3/3 genotype (genotype which is less prevalent among IAA+ children). |

Supplemental Table 1. Autoantibody features characterize progression before T1D diagnosis.

| | | | | | | |
|---|---|---|---|---|---|---|
| Krischer 2017 (TEDDY)[42] | 8,503 | Pediatric | • High genetic risk | • Aab type | Yes | • Differences in autoimmunity initiation according to genetic and environmental factors drive either GAD or IAA as first appearing Aab. |
| Pöllänen 2017 (DIPP)[43] | 7,410 | Pediatric | • High genetic risk | • Aab number<br>• Aab titer<br>• Aab type<br>• Aab timing<br>• Age at Aab seroconversion | Yes | • Compared with slower progressors to T1D, rapid progressors had a higher frequency of positivity for multiple Aabs and had higher titers of ICA, IAA and IA-2 at seroconversion.<br>• Compared with Aab+ non-progressors, rapid progressors were younger, were more likely to carry the high-risk HLA genotype, have multiple Aabs, predisposing SNP in the PTPN22 gene, higher frequency of ICA, IAA, GAD and IA-2, and higher titers of all four autoantibodies at seroconversion. |

Supplemental Table 1. Autoantibody features characterize progression before T1D diagnosis.

| | | | | | | |
|---|---|---|---|---|---|---|
| Sosenko 2017 (TrialNet)[44] | 272 | Pediatric and adult | • FDR<br>• Single Aab+<br>• Other: single GAD or IAA+ | • Aab number<br>• Aab affinity<br>• Aab type<br>• Other: ECL as measure of Aab affinity | No | • Progression to multiple Aabs was higher for GAD/ECL+ than GAD/ECL-.<br>• Progression to multiple Aabs was higher for and IAA/ECL+ than IAA/ECL-.<br>• ECL measurements appear to have utility for natural history studies and prevention trials of individuals with single Aabs.<br>• ECL+ are at appreciable risk for developing multiple Aabs and for glycemic progression toward T1D; ECL-are at very low risk. |
| Strollo 2017 (ABIS)[45] | 86 | Pediatric | • Single Aab+<br>• Multiple Aab+<br>• Other: general population controls | • Aab type<br>• Novel Aab/epitope | Not available | • At least one oxPTM-INS Ab were present 91.3% of progressors to T1D.<br>• OH-INS-Ab were more common in progressors vs nonprogressors (82.6% vs 19%) and allowed discrimination between progressors and nonprogressors with 74% sensitivity and 91% specificity. |
| Viisanen 2017 (DIPP)[46] | 276 | Pediatric | • High genetic risk<br>• Single Aab+<br>• Multiple Aab+<br>• New onset | • Aab number<br>• Aab type | No | • Frequency of CXCR5+PD-1+ICOS+ activated Tfh cells is increased in children with new onset T1D and multiple ab+ children with impaired glucose tolerance.<br>• No alterations in circulating B cell compartments before or after T1D onset. |
| Balke 2018 (BDR)[47] | 461 | Pediatric and adult | • FDR<br>• Single Aab+<br>• Multiple Aab+ | • Aab number<br>• Aab type<br>• Aab timing | Yes | • Progression from single Aab+ to clinical onset not influenced by HLA-A*24, -B*18, or -B*39 status<br>• Progression from single to multiple Aab+ was delayed in the presence of HLA-A*24 but not in the presence of HLA-B*18 or -B*39 (independent of older age and absence of HLA-DQ2/DQ8 or -DQ8, and only in the presence of GAD Aab).<br>• HLA-A*24 was associated with accelerated progression from multiple Aab+ to T1D but its effects were restricted to HLA-DQ8+, IA-2+ or ZnT8+ relatives.<br>• HLA-B*18, but not -B*39, was associated with more rapid progression, but only in HLA-DQ2 carriers with double positivity for GAD and IAA |

Supplemental Table 1. Autoantibody features characterize progression before T1D diagnosis.

| | | | | | | |
|---|---|---|---|---|---|---|
| Ilonen 2018 (DIPP)[48] | 128 | Pediatric | • High genetic risk<br>• Single Aab+ | • Aab type<br>• Age at Aab seroconversion<br>• Order of Aab seroconversion | Yes | • In children at high genetic risk followed from birth, most common primary antibodies were IAA, followed by GAD, IA-2, and ZnT8.<br>• In many cases, IAA had disappeared by diagnosis. |
| Long 2018 (BABYDIAB, DAISY, ABIS, BOX and Pittsburgh)[49] | 132 | Pediatric and adult | • High genetic risk<br>• FDR<br>• Multiple Aab+ | • Aab number<br>• Aab type | Yes | • In slow progressors, the most frequent Aabs were GAD (92%), followed by ZnT8 (62%), IAA (59%) and IA-2 (41%).<br>• Four major autoantibodies (IAA, GAD, IA-2 and ZnT8) were all detected in slow progressors. |
| Redondo 2018 (TrialNet)[50] | 244 | Pediatric and adult | • FDR<br>• Second degree relative<br>• Single Aab+<br>• Other: participants with Immunochip data | • Aab number<br>• Aab type<br>• Aab timing | No | • In subjects with family history of T1D, the TCF7L2 locus did not significantly predict progression to multiple Aab+.<br>• Among single GAD+ participants, those who carried at least one TCF7L2 allele had a lower rate of progression to multiple Aab+ than those who did not, after adjustment for HLA risk haplotypes and age.<br>• Among subjects who were either IA-2 or insulin autoantibody positive only, carrying at least one TCF7L2 risk allele was not a significant factor overall, but in overweight or obese participants, it increased the risk of progression to multiple autoantibody positivity even with adjustment for age. |
| Redondo 2018 (TrialNet)[51] | 1,244 | Pediatric and adult | • FDR<br>• Second degree relative<br>• Single Aab+<br>• Multiple Aab+ | • Aab number<br>• Aab type | Yes | • Higher GRS was significantly associated with increased progression rate from single to multiple positive Aabs after adjusting for age, autoantibody type, ethnicity, and sex.<br>• Progression to T1D was best predicted by a combined model with GRS, number of positive autoantibodies, DPT-1 Risk Score, and age. |
| Sanda 2018 (TrialNet)[52] | 778 | Pediatric and adult | • Multiple Aab+ | • Aab titer<br>• Aab type | Not available | • Higher cumulative titer both ICA and GAD correlated with greater incident dysglycemia (i.e. an impaired or diabetes range OGTT)<br>• Both cumulative mean titer levels of ICA and longitudinal increases in ICA titer levels correlate with abnormal OGTT results in individuals at risk for T1D. |

Supplemental Table 1. Autoantibody features characterize progression before T1D diagnosis.

| | | | | | | |
|---|---|---|---|---|---|---|
| | | | | | | • Fluctuations in Aab titers do not correlate with lower rates of progression to clinical disease. |
| Sharma 2018 (TEDDY)[53] | 5,806 | Pediatric | • High genetic risk | • Aab number<br>• Aab type<br>• Age at Aab seroconversion<br>• Order of Aab seroconversion | Not available | • Novel gene region PPIL2 was associated with time to any persistent confirmed islet Aab and time to T1D.<br>• Gene region INS/TH is associated with time to multiple Aab, time to first appearing Aab, and time to T1D.<br>• Known region PTPN22 was associated with time to any persistent confirmed islet Aab and time to multiple Aab but not time to T1D.<br>• Known region SH2B3 was associated with time to any persistent confirmed islet Aab but not time to T1D.<br>• Novel regions RNASET2/MIR3939 and PLEKHA1/MIR3941 were found to be associated with time to T1D but they were not associated with time to persistent confirmed islet Aab, time to multiple Aab, time to IAA as the first appearing Aab, or time to GAD as the first appearing Aab.<br>• Novel region RBFOX1 was associated with time to GAD as the first appearing Aab but not time to T1D.<br>• Novel region TTC34 was associated with time to IAA as the first appearing Aab but not time to T1D.<br>• Novel region PXK/PDHB was associated with time to multiple Aab but not time to T1D. |
| Sioofy-Khojine 2018 (DIPP)[54] | 501 | Pediatric | • High genetic risk<br>• Other: single IAA or GAD+ but later converted to multiple Aab+ | • Aab number<br>• Aab type<br>• Order of Aab seroconversion | No | • There is an association between CVB1infection and progression to multiple Aab and T1D in individuals who develop IAA as a first single Aab.<br>• Infection of viruses CVB[2-6] were not associated with T1D development in children who develop IAA as a first single Aab.<br>• Infection of viruses CVB[1-6] were not associated with T1D development in children who developed single Aab positivity with GAD first. |

Supplemental Table 1. Autoantibody features characterize progression before T1D diagnosis.

| | | | | | | |
|---|---|---|---|---|---|---|
| Steck 2018 (DAISY)[55] | 68 | Pediatric | • High genetic risk<br>• FDR<br>• Other: General population with at least one Aab | • Aab number<br>• Aab titer<br>• Aab type<br>• Aab timing<br>• Age at Aab seroconversion | Yes | • Age at seroconversion, number of Aabs, IA-2 levels, HbA1c and metabolic variables from the OGTT predicted progression to diabetes.<br>• A model containing age at seroconversion, number of Ab+, IA-2 levels, HbA1c, 1h glucose and 1h C-peptide was as predictive for T1D as progression as models including all sum or AUC values for glucose and C-peptide from full OGTT. |
| Acevedo-Calado 2019 (TrialNet)[56] | 1,686 | Pediatric and adult | • FDR<br>• Single Aab+<br>• Multiple Aab+ | • Novel Aab/epitope | Yes | • Identified AAbs reacting with a variant IA-2 molecule (IA-2var).<br>• IA-2var+ FDRs with negative classic IA-2 testing but GAD and/or IAA+ had faster progression to T1D than IA-2var- FDRs.<br>• Single Aab+ FDRs with HLADR*04-DQB1*03:02 and IA-2var+ showed rapid progression to T1D. |
| Bauer 2019 (DIPP)[57] | 15,253 | Pediatric | • High genetic risk | • Age at Aab seroconversion<br>• Order of Aab seroconversion | Yes | • Young age at initial seroconversion was associated with a high probability of IAA-initiated autoimmunity and progression to T1D.<br>• GAD-initiated autoimmunity and progression to diabetes were not dependent on initial seroconversion age.<br>• Strength of HLA risk affected the progression of both IAA and GAD initiated autoimmunity<br>• The simultaneous appearance of two other Aabs increased the rate of progression to T1D compared with that of a single secondary Aab among subjects with GAD but not IAA initiated autoimmunity. |
| Beyerlein 2019 (TEDDY)[58] | 341 | Pediatric | • High genetic risk<br>• Single Aab+<br>• Multiple Aab+ | • Age at Aab seroconversion | Yes | • Age of Aab development (either seroconversion or multiple Aab) associated with faster progression to next stage (multiple Aab or stage 3).<br>• Age at seroconversion was significant component of model for association of genetic risk score with progression. |
| Endesfelder 2019 (TEDDY)[59] | 600 | Pediatric | • High genetic risk | • Aab number<br>• Aab titer<br>• Aab type<br>• Aab timing | Yes | • Early and stable seroconversion to IAA and IA-2 conferred high progression risk that was unaffected by GAD status. |

Supplemental Table 1. Autoantibody features characterize progression before T1D diagnosis.

| | | | | | | |
|---|---|---|---|---|---|---|
| | | | | • Age at Aab seroconversion<br>• Order of Aab seroconversion | | |
| Jacobsen 2019 (TEDDY)[60] | 363 | Pediatric | • High genetic risk<br>• Single Aab+<br>• Multiple Aab+ | • Aab number<br>• Aab titer<br>• Aab type<br>• Age at Aab seroconversion | Yes | • Logistic regression modelling identified 5 significant predictors of progression to T1D: IA-2 status, HbA1c, BMI Z-score, SNP rs1270876_G, and a combination marker of Aab number plus fasting insulin level. |
| Krischer 2019 (TEDDY)[61] | 7,777 | Pediatric | • High genetic risk<br>• Single Aab+<br>• Multiple Aab+<br>• New onset | • Aab number<br>• Aab type<br>• Aab timing<br>• Age at Aab seroconversion<br>• Order of Aab seroconversion | Yes | • HLA genotype (DR3/4 vs. others) was the best predictor for islet autoimmunity.<br>• PTPN22 rs2476601 was the best predictor for IAA+ first.<br>• Weight at 1 year was the best predictor for GAD Aab appearing first.<br>• In a multivariate model, IAA first was best predictor of T1D at 6 years.<br>• AUC of the prediction model for T1D at 3 years after the appearance of multiple Aab reached 0.706 (95% CI 0.649, 0.762). |
| Paun 2019 (TrialNet)[62] | 130 | Pediatric | • FDR<br>• Single Aab+<br>• Multiple Aab+<br>• New onset | • Aab type | Yes | • Serum IgG2 to roseburia faecis and bacterial consortium were associated with progression to T1D in multiple Aab+ FDRs with DR3/4 haplotypes. |
| Pöllänen 2019 (DIPP)[63] | 7,410 | Pediatric | • High genetic risk | • Aab titer<br>• Aab type<br>• Age at Aab seroconversion<br>• Order of Aab seroconversion | Yes | • Compared with other progressors, slow progressors were less often multiple Aab+, had lower ICA and IAA titers, and lower frequency of IA-2 at seroconversion.<br>• Compared with multiple Aab+ nonprogressors, slow progressors had higher ICA titers, and higher frequency of IAA and multiple Aabs at seroconversion.<br>• Multiple Aab+ nonprogressors were more often GAD+ without IAA at seroconversion. |
| Simmons 2019 (TrialNet)[64] | 57 | Pediatric and adult | • FDR<br>• Second degree relative<br>• Single Aab+ | • Aab number<br>• Aab type | Yes | • Close to T1D (24 mo prior) onset, unmethylated INS ratio associated with IAA, ECL-IAA, and IA-2 levels, but not GAD, ECL-GAD, ECL-IA-2 or ZnT8. |

Supplemental Table 1. Autoantibody features characterize progression before T1D diagnosis.

| | | | | | | |
|---|---|---|---|---|---|---|
| | | | • Multiple Aab+ | | | • Only IAA levels associated with unmethylated INS ratio over time. |
| Strollo 2019 (ABIS)[65] | 55 | Pediatric | • Single Aab+<br>• Multiple Aab+ | • Aab type<br>• Novel Aab/epitope | No | • Aabs to posttranslationally modified, oxidized insulin (oxPTM-INS-Ab) improve T1D risk assessment and prediction accuracy in Aab+ children.<br>• oxPTM-INS-Ab prediction accuracy is comparable to IA-2A and higher than GAD and IAA. |
| Triolo 2019 (TrialNet)[66] | 17,226 | Pediatric and adult | • FDR<br>• Single Aab+<br>• Other: twins and siblings of individuals with T1D | • Aab type<br>• Aab timing | Yes | • Identical twins were more likely to be GAD, IA-2, and/or IAA+ than non-identical twins or full siblings.<br>• In initially Aab- subjects, 1.5% of identical twins, 0% of nonidentical twins, and 0.5% of full siblings progressed to diabetes at 3 years of follow-up.<br>• For initially single Aab+ subjects, at 3 years of follow-up, 69% of identical twins, 13%of nonidentical twins, and 12% of full siblings developed type 1 diabetes. |
| Ferrat 2020 (TEDDY)[67] | 7,798 | Pediatric | • High genetic risk | • Aab titer<br>• Aab type<br>• Aab timing<br>• Age at Aab seroconversion<br>• Other: presence of persistent islet autoantibodies (GADA, IA2A, IAA) | Yes | • Combined Risk Score (CRS) with genetic, clinical and immunological data improves T1D prediction. up to 8 years of age compared with Aabs alone.<br>• CRS helped most for Aab- or single Aab+ children. |
| Hanna 2020 (BOX)[68] | 69 | Adult | • FDR<br>• Multiple Aab+<br>• Other: "slow progressors" multiple Aab+ and T1D free after 10 years | • Aab number<br>• Aab titer<br>• Aab type<br>• Other: Aab reversion | Not available | • Slow progressors lose islet Aabs over time and generally the titers decrease as well.<br>• Presence of Aabs in the serum may be due to lower thresholds of B cell stimulation for antibody production in the slow progressors. |
| Jacobsen 2020 (TrialNet)[69] | 1,815 | Pediatric | • FDR<br>• Second degree relative | • Aab number<br>• Aab type | Yes | • Risk of T1D progression was variable in multiple Aab+ children depending on: age (inversely |

Supplemental Table 1. Autoantibody features characterize progression before T1D diagnosis.

| | | | | | | |
|---|---|---|---|---|---|---|
| | | | • Multiple Aab+ | | | related); GAD (less risk); IA-2 (higher risk); low Index60 (lower risk). |
| Li 2020 (TEDDY)[70] | 1,648 | Pediatric | • High genetic risk<br>• FDR | • Aab type<br>• Aab timing<br>• Age at Aab seroconversion<br>• Order of Aab seroconversion | Yes | • Differences in metabolite levels seen prior to development of IAA as first Aab vs GAD as a first Aab. |
| Liu 2020 (TEDDY)[71] | 7,522 | Pediatric | • High genetic risk<br>• FDR | • Aab type<br>• Aab timing<br>• Age at Aab seroconversion<br>• Order of Aab seroconversion | Yes | • Higher rate of weight gain in early childhood was associated with increased risk of progression from islet autoimmunity to T1D in children with first-appearing GAD Aab only.<br>• Lower height growth rate in infancy and a higher height growth rate in early childhood appeared to be more associated with the progression from islet autoimmunity to T1D in children who had GAD only or multiple Aabs at initial seroconversion. |
| Mikk 2020 (DIPP, FPDR)[72] | 621 | Pediatric | • Single Aab+ | • Aab number<br>• Aab type<br>• Age at Aab seroconversion | Not available | • Homozygotes for the DR3-DQ2 haplotype had almost exclusively GAD as the first Aab.<br>• More even distribution between GAD and IAA was found in DR3-DQ2/DR4-DQ8 and DR3-DQ/x and DR4-DQ8/x genotypes.<br>• In DR4-DQ8 positive genotypes with the DRB1*04:01 allele IAA was more often the first autoantibody than in DRB1*04:04 positive genotypes. |
| Pöllänen 2020 (DIPP)[73] | 1,006 | Pediatric | • High genetic risk | • Aab number<br>• Aab titer<br>• Aab type<br>• Aab timing<br>• Age at Aab seroconversion<br>• Order of Aab seroconversion | Yes | • In HLA predisposed children, the primary Aab is characteristic of age (< 2 years, ZnT8 and IAA most commonly presented first, IA-2 and GAD autoimmunity increased in preschool age, and GAD+ seroconversions continued to appear steadily until age 10-15 years).<br>• Inverse IAA seroconversions (positive to negative) occurred frequently and marked a prolonged delay from seroconversion to diagnosis compared to persistent IAA. |
| So 2020 (TrialNet)[74] | 3,284 | Pediatric and adult | • FDR | • Aab number<br>• Aab titer | Yes | • Of 3,284 multiple Aab+ subjects, reversion occurred in 134 (4.1%). |

Supplemental Table 1. Autoantibody features characterize progression before T1D diagnosis.

| | | | | | | |
|---|---|---|---|---|---|---|
| | | | • Second degree relative<br>• Multiple Aab+ | • Aab type<br>• Aab timing<br>• Order of Aab seroconversion<br>• Other: Aab reversion | | • Reversion was associated with reduced 5 year incidence of clinical disease.<br>• Reversion occurred more frequently with older age.<br>• Reversion occurred more frequently if lower autoantibody titers.<br>• Reversion occurred more frequently if fewer positive Aabs. |
| Vehik 2020 (TEDDY)[75] | 608 | Pediatric | • High genetic risk<br>• FDR | • Aab number<br>• Aab type<br>• Aab timing<br>• Age at Aab seroconversion<br>• Order of aab seroconversion | Yes | • Risk of second Aab was independent of GAD vs. IAA as first Aab.<br>• Second appearing GAD, IAA, IA-2, or ZnT8 conferred increased risk for T1D compared to children who maintained single Aab+.<br>• If second appearing Aab was IA-2, greater risk of disease progression compared to GAD or IAA.<br>• Younger age at seroconversion and shorter time to development of second Aab increased T1D risk. |
| Baumann 2021 (Karlsburg Type 1 Diabetes Risk Study)[76] | 239 | Pediatric and adult | • Single Aab+<br>• Multiple Aab+<br>• New onset | • Aab number<br>• Aab type | No | • ZnT8 Aab+ contributed to risk stratification in IA-2 Aab+ schoolchildren.<br>• ZnT8 Aab+ identified individuals with disease progression who were negative for IA-2.<br>• Associations with GAD and IAA were not identified. |
| Bediaga 2021 (TrialNet, DPT-1, TEDDY, Fr1da)[77] | 2,973 | Pediatric and adult | • High genetic risk<br>• FDR<br>• Second degree relative<br>• Single Aab+<br>• Multiple Aab+ | • Aab number<br>• Aab titer<br>• Aab type<br>• Age at Aab seroconversion | Yes | • The single time point risk scores M60 and M120 predict progression from stage1/2 to stage 3 T1D. |
| Bonifacio 2021 (TEDDY)[78] | 8,556 | Pediatric | • High genetic risk | • Aab number<br>• Aab type<br>• Age at Aab seroconversion | Yes | • Risk of developing islet autoimmunity declines exponentially with age.<br>• Major influence of genetic factors is limited to the first few years of life.<br>• Highest sensitivity and positive predictive value of multiple islet autoantibody phenotypes for T1D was achieved by Aab screening at 2 yrs and again at 5–7 yrs of age. |

Supplemental Table 1. Autoantibody features characterize progression before T1D diagnosis.

| | | | | | | |
|---|---|---|---|---|---|---|
| Jia 2021 (ASK)[79] | 135 | Pediatric | • Single Aab+<br>• Multiple Aab+<br>• New onset | • Comparison of Aab assays | Not available | • In Aab+ children, ZnT8 positivity on ECL assay was associated with more rapid progression to disease than RBA positivity.<br>• Positive predictive value of ZnT8 ECL was significantly higher than RBA. |
| Jia 2021 (DAISY)[79] | 123 | Pediatric | • High genetic risk<br>• FDR<br>• Single Aab+<br>• Multiple Aab+ | • Comparison of Aab assays | Not available | • Of 11 children positive for ZnT8 only by RBA, 3 were also positive for ZnT8 by ECL and all 3 progressed to T1D. |
| Korneva 2021[80] | 424 | Pediatric | • FDR<br>• Single Aab+<br>• Multiple Aab+<br>• New onset | • Aab number<br>• Aab titer<br>• Aab type<br>• Aab timing<br>• Age at Aab seroconversion | Yes | • Diabetes progression in children with a sibling with T1DM was most common in the individuals with at least 2 AAbs, within 1-2 years of single AAb appearance.<br>• IA-2 and ZnT8 appeared later (with more rapid progression of the disease).<br>• IAA+ was more common in younger children. |
| Krischer 2021 (TEDDY)[81] | 8,502 | Pediatric | • High genetic risk<br>• Single Aab+<br>• Multiple Aab+ | • Aab number<br>• Aab titer<br>• Aab type<br>• Aab timing<br>• Age at Aab seroconversion<br>• Order of aab seroconversion | Yes | • Children who progressed to T1D prior to age 6 developed Aabs earlier and progressed to T1D more rapidly than children who were diagnosed between 6-13 years of age.<br>• IAA appeared before GAD.<br>• IA-2 appeared first only in the older (6-13 year at onset age) group.<br>• Children who seroconverted to multiple Aabs after age 2 had lower risk than those who converted before age 2; risk did not further decrease after age 2. |
| Long 2021[82] | 221 | Pediatric | • Single Aab+<br>• Multiple Aab+ | • Aab number<br>• Aab type<br>• Order of aab seroconversion<br>• Novel Aab/epitope | Not available | • Autoantibody profiles differ between Lithuanian compared with English schoolchildren, which may help to inform which primary screening to obtain in these populations. |
| Nevalainen 2021 (DIPP)[83] | 6,081 | Pediatric | • High genetic risk | • Aab number<br>• Aab type<br>• Age at Aab seroconversion | | • High genetic risk is associated with the appearance of all Aabs.<br>• Association of sex and urban municipality associated with IA-2 and IAA only. |

Supplemental Table 1. Autoantibody features characterize progression before T1D diagnosis.

| | | | | | | |
|---|---|---|---|---|---|---|
| Vandewalle 2021 (BDR)[84] | 462 | Pediatric and adult | • FDR<br>• Second degree relative | • Aab number<br>• Aab type<br>• Aab timing<br>• Age at Aab seroconversion | Not available | • Progression from single to multiple Aab+ was delayed in females with ERBB3 GG or IKZF4 TT genotypes, but not in males.<br>• Progression from multiple Aab+ to T1D was not influenced by ERBB3/IKZF4. |
| Kwon 2022 (DAISY, DiPiS, DIPP, DEW-IT, BABYDIAB)[85] | 2,172 | Pediatric and adult | • High genetic risk<br>• FDR<br>• Single Aab+<br>• Multiple Aab+ | • Aab number<br>• Aab type<br>• Aab timing<br>• Order of Aab seroconversion | Yes | • Patterns of AAb sequences predict rates of progression to clinical (stage 3) T1D (3 "trajectories").<br>• Age, sex, and HLA-DR status further refine the progression rates within trajectories. |
| Li 2022 (T1DI)[86] | 10, 145 | Pediatric | • High genetic risk<br>• FDR | • Aab type<br>• Age at Aab seroconversion | Yes | • Height rate was positively associated with development of T1D in the analyses from seroconversion with IAA to T1D.<br>• Rapid increase in height (cm/year) was associated with increased risk of seroconversion to GAD, IAA, or IA-2 for 1-3 years of age (HR 1.26) and >3 years of age (HR 1.48). |
| Martinez 2022 (TEDDY)[87] | 57 | Pediatric and adult | • High genetic risk<br>• FDR<br>• Multiple Aab+ | • Aab number<br>• Aab type | Yes | • No association between the number or types of Aabs and baseline measures of glucose metabolism and beta cell function in multiple Aab+ individuals.<br>• The presence of ZnT8(Q/R/W)A was associated with lower first phase insulin response. |
| Ng 2022 (T1DI)[88] | 1,604 | Pediatric | • High genetic risk<br>• FDR<br>• Single Aab+ | • Aab titer | Yes | • Minimum titer associated with a maximum difference in 5-year risk differed for each Aab when stratified by quartiles of titer.<br>• Aab type specific titer thresholds identified children with a greater than or equal to 50% 5-year risk when considering age-specific Aab screening. |
| So 2022 (TrialNet)[89] | 8,749 | Pediatric and adult | • FDR<br>• Second degree relative<br>• Other: third degree relative | • Aab number<br>• Aab type<br>• Age at Aab seroconversion | Yes | • In single Aab+ subjects, DR3, GAD+, elevated BMI, and HOMA-IR showed increase in effect on progression with increasing age.<br>• IAA had a diminishing effect with older age on progression in single Aab+.<br>• In multiple Aab+, male sex was associated with increased risk for progression and DR3/4 showed decreased effect with older age. |

Supplemental Table 1. Autoantibody features characterize progression before T1D diagnosis.

| | | | | | | |
|---|---|---|---|---|---|---|
| | | | | | | • Index60 was a better predictor in young vs old single and multiple Aab+ subjects.<br>• Age did not discriminate effectiveness of DPTRS. |

*Participant groups were considered new onset if within 12 months of T1D diagnosis

Abbreviations: Aab (autoantibody), ABIS (All Babies in Southwest Sweden), ASK (Autoimmunity Screening in Kids), BABYDIAB and BABYDIET (German longitudinal birth cohort studies), BOX (Bart's Oxford Family Study), BDR (Belgian Diabetes Registry), DAISY (Diabetes Autoimmunity Study in the Young), DEW-IT (Diabetes Evaluation in Washington), DiAPREV-IT (Diabetes Prevention-Immune Tolerance), DIPP (Finnish Type 1 Diabetes Prediction and Prevention Project), DiPiS (Diabetes Prediction in Skåne), DPT-1 (Diabetes Prevention Trial Type 1), DPTRS (Diabetes Prevention Trial-Type 1 Risk Score), ECL (electrochemiluminescent), ENDIT (European Nicotinamide Diabetes Intervention Trial), FDR (first degree relative), FPDR (Finnish Pediatric Diabetes Register), Fr1da (Early Detection for Early Care of Type 1 Diabetes), GAD (glutamic acid decarboxylase antibody), HOMA-IR (homeostatic model assessment of insulin resistance), IA-2 (islet antigen-2 antibody), IAA (insulin autoantibody), ICA (islet cell autoantibody), OGTT (oral glucose tolerance test), RBA (radio binding assay), T1DI (Type 1 Diabetes Intelligence), TEDDY (The Environmental Determinants of Diabetes in the Young), Tfh (T follicular helper), ZnT8 (zinc transporter antibody)

Supplemental Table 1. Autoantibody features characterize progression before T1D diagnosis.

34. Steck, A. K. *et al.* ECL-IAA and ECL-GADA Can Identify High-Risk Single Autoantibody-Positive Relatives in the TrialNet Pathway to Prevention Study. *Diabetes Technol. Ther.* **18**, 410–414 (2016).
35. Vehik, K. *et al.* Reversion of β-Cell Autoimmunity Changes Risk of Type 1 Diabetes: TEDDY Study. *Diabetes Care* **39**, 1535–1542 (2016).
36. Xu, P. & Krischer, J. P. Prognostic Classification Factors Associated With Development of Multiple Autoantibodies, Dysglycemia, and Type 1 Diabetes-A Recursive Partitioning Analysis. *Diabetes Care* **39**, 1036–1044 (2016).
37. Bosi, E. *et al.* Impact of Age and Antibody Type on Progression From Single to Multiple Autoantibodies in Type 1 Diabetes Relatives. *J. Clin. Endocrinol. Metab.* **102**, 2881–2886 (2017).
38. Frohnert, B. I. *et al.* Late-onset islet autoimmunity in childhood: the Diabetes Autoimmunity Study in the Young (DAISY). *Diabetologia* **60**, 998–1006 (2017).
39. Gorus, F. K. *et al.* Twenty-Year Progression Rate to Clinical Onset According to Autoantibody Profile, Age, and HLA-DQ Genotype in a Registry-Based Group of Children and Adults With a First-Degree Relative With Type 1 Diabetes. *Diabetes Care* **40**, 1065–1072 (2017).
40. Krischer, J. P. *et al.* The Influence of Type 1 Diabetes Genetic Susceptibility Regions, Age, Sex, and Family History on the Progression From Multiple Autoantibodies to Type 1 Diabetes: A TEDDY Study Report. *Diabetes* **66**, 3122–3129 (2017).
41. Köhler, M. *et al.* Joint modeling of longitudinal autoantibody patterns and progression to type 1 diabetes: results from the TEDDY study. *Acta Diabetol.* **54**, 1009–1017 (2017).
42. Krischer, J. P. *et al.* Genetic and Environmental Interactions Modify the Risk of Diabetes-Related Autoimmunity by 6 Years of Age: The TEDDY Study. *Diabetes Care* **40**, 1194–1202 (2017).
43. Pöllänen, P. M. *et al.* Characterisation of rapid progressors to type 1 diabetes among children with HLA-conferred disease susceptibility. *Diabetologia* **60**, 1284–1293 (2017).
44. Sosenko, J. M. *et al.* The Use of Electrochemiluminescence Assays to Predict Autoantibody and Glycemic Progression Toward Type 1 Diabetes in Individuals with Single Autoantibodies. *Diabetes Technol. Ther.* **19**, 183–187 (2017).
45. Strollo, R. *et al.* Antibodies to post-translationally modified insulin as a novel biomarker for prediction of type 1 diabetes in children. *Diabetologia* **60**, 1467–1474 (2017).
46. Viisanen, T. *et al.* Circulating CXCR5+PD-1+ICOS+ Follicular T Helper Cells Are Increased Close to the Diagnosis of Type 1 Diabetes in Children With Multiple Autoantibodies. *Diabetes* **66**, 437–447 (2017).
47. Balke, E. M. *et al.* Accelerated Progression to Type 1 Diabetes in the Presence of HLA-A*24 and -B*18 Is Restricted to Multiple Islet Autoantibody-Positive Individuals With Distinct HLA-DQ and Autoantibody Risk Profiles. *Diabetes Care* **41**, 1076–1083 (2018).
48. Ilonen, J. *et al.* Primary islet autoantibody at initial seroconversion and autoantibodies at diagnosis of type 1 diabetes as markers of disease heterogeneity. *Pediatr. Diabetes* **19**, 284–292 (2018).
49. Long, A. E. *et al.* Characteristics of slow progression to diabetes in multiple islet autoantibody-positive individuals from five longitudinal cohorts: the SNAIL study. *Diabetologia* **61**, 1484–1490 (2018).
50. Redondo, M. J. *et al.* TCF7L2 Genetic Variants Contribute to Phenotypic Heterogeneity of Type 1 Diabetes. *Diabetes Care* **41**, 311–317 (2018).
51. Redondo, M. J. *et al.* A Type 1 Diabetes Genetic Risk Score Predicts Progression of Islet Autoimmunity and Development of Type 1 Diabetes in Individuals at Risk. *Diabetes Care* **41**, 1887–1894 (2018).
52. Sanda, S. Increasing ICA512 autoantibody titers predict development of abnormal oral glucose tolerance tests. *Pediatr. Diabetes* **19**, 271–276 (2018).
53. Sharma, A. *et al.* Identification of non-HLA genes associated with development of islet autoimmunity and type 1 diabetes in the prospective TEDDY cohort. *J. Autoimmun.* **89**, 90–100 (2018).
54. Sioofy-Khojine, A. B. *et al.* Coxsackievirus B1 infections are associated with the initiation of insulin-driven autoimmunity that progresses to

type 1 diabetes. *Diabetologia* **61**, 1193–1202 (2018).
55. Steck, A. K. *et al.* Predicting progression to diabetes in islet autoantibody positive children. *J. Autoimmun.* **90**, 59–63 (2018).
56. Acevedo-Calado, M. J. *et al.* Autoantibodies Directed Toward a Novel IA-2 Variant Protein Enhance Prediction of Type 1 Diabetes. *Diabetes* **68**, 1819–1829 (2019).
57. Bauer, W. *et al.* Age at Seroconversion, HLA Genotype, and Specificity of Autoantibodies in Progression of Islet Autoimmunity in Childhood. *J. Clin. Endocrinol. Metab.* **104**, 4521–4530 (2019).
58. Beyerlein, A. *et al.* Progression from islet autoimmunity to clinical type 1 diabetes is influenced by genetic factors: results from the prospective TEDDY study. *J. Med. Genet.* **56**, 602–605 (2019).
59. Endesfelder, D. *et al.* Time-Resolved Autoantibody Profiling Facilitates Stratification of Preclinical Type 1 Diabetes in Children. *Diabetes* **68**, 119–130 (2019).
60. Jacobsen, L. M. *et al.* Predicting progression to type 1 diabetes from ages 3 to 6 in islet autoantibody positive TEDDY children. *Pediatr. Diabetes* **20**, 263–270 (2019).
61. Krischer, J. P. *et al.* Predicting islet cell autoimmunity and type 1 diabetes: An 8-year teddy study progress report. *Diabetes Care* **42**, 1051–1060 (2019).
62. Paun, A. *et al.* Association of HLA-dependent islet autoimmunity with systemic antibody responses to intestinal commensal bacteria in children. *Sci. Immunol.* **4**, (2019).
63. Pöllänen, P. M. *et al.* Characteristics of Slow Progression to Type 1 Diabetes in Children With Increased HLA-Conferred Disease Risk. *J. Clin. Endocrinol. Metab.* **104**, 5585–5594 (2019).
64. Simmons, K. M. *et al.* Unmethylated Insulin as an Adjunctive Marker of Beta Cell Death and Progression to Type 1 Diabetes in Participants at Risk for Diabetes. *Int. J. Mol. Sci.* **20**, (2019).
65. Strollo, R. *et al.* Antibodies to oxidized insulin improve prediction of type 1 diabetes in children with positive standard islet autoantibodies. *Diabetes. Metab. Res. Rev.* **35**, (2019).
66. Triolo, T. M. *et al.* Identical and Nonidentical Twins: Risk and Factors Involved in Development of Islet Autoimmunity and Type 1 Diabetes. *Diabetes Care* **42**, 192–199 (2019).
67. Ferrat, L. A. *et al.* A combined risk score enhances prediction of type 1 diabetes among susceptible children. *Nat. Med.* **26**, 1247–1255 (2020).
68. Hanna, S. J. *et al.* Slow progressors to type 1 diabetes lose islet autoantibodies over time, have few islet antigen-specific CD8+ T cells and exhibit a distinct CD95hi B cell phenotype. *Diabetologia* **63**, 1174–1185 (2020).
69. Jacobsen, L. M. *et al.* The risk of progression to type 1 diabetes is highly variable in individuals with mulitple autoantibodies following screening. *Diabetologia* **63**, 588–596 (2020).
70. Li, Q. *et al.* Longitudinal Metabolome-Wide Signals Prior to the Appearance of a First Islet Autoantibody in Children Participating in the TEDDY Study. *Diabetes* **69**, 465–476 (2020).
71. Liu, X. *et al.* Distinct Growth Phases in Early Life Associated With the Risk of Type 1 Diabetes: The TEDDY Study. *Diabetes Care* **43**, 556–562 (2020).
72. Mikk, M. L. *et al.* HLA-DR-DQ haplotypes and specificity of the initial autoantibody in islet specific autoimmunity. *Pediatr. Diabetes* **21**, 1218–1226 (2020).
73. Pöllänen, P. M. *et al.* Dynamics of Islet Autoantibodies During Prospective Follow-Up From Birth to Age 15 Years. *J. Clin. Endocrinol. Metab.* **105**, (2020).
74. So, M., O'Rourke, C., Bahnson, H. T., Greenbaum, C. J. & Speake, C. Autoantibody Reversion: Changing Risk Categories in Multiple-Autoantibody-Positive Individuals. *Diabetes Care* **43**, 913–917 (2020).

Supplemental Table 2. Autoantibody features characterize heterogeneity at T1D diagnosis.

| Study | n | Age group | Population Studied* | Autoantibody Feature Assessed | Age impact? | Findings |
|---|---|---|---|---|---|---|
| Andersson 2011[1] | 686 | Pediatric | • New onset | • Aab number<br>• Aab type | Yes | • Aab- T1D was more common in older children.<br>• ZnT8 associated with DQ-A1-B1*X-0604 and DQ-A1-B1*X-04.<br>• ICA associated with DQA1-B1*03-0302.<br>• GAD positively and IA-2A negatively associated with DQA1-B1*05-02. |
| Brorsson 2011 (DSBD)[2] | 960 | Pediatric | • Aab+ siblings<br>• New onset | • Aab number<br>• Aab titer<br>• Aab type<br>• Novel Aab/ epitope | Yes | • Positive correlation between rs13266634 genotype and specificity for the ZnT8-Arginine (ZnT8R) and ZnT8-Tryptophan (ZnT8W) isoforms.<br>• Significant correlation between ZnT8R+ and HLA-DQB1*0302 genotype.<br>• ZnT8A+ overlapped substantially with GAD+ and IA-2+ and correlated with IA-2+.<br>• ZnT8A+ did not affect insulin dose-adjusted HbA1c.<br>• Lower ZnT8R+ and GAD+ prevalence and titers found in probands with age of diagnosis <5 yrs. |
| Howson 2011 (ADBW-END)[3] | 1,384 | Pediatric and adult | • New onset | • Aab type | | • DR3 was associated with GAD+ but IA-2-.<br>• DR4 was associated with IA-2+. |
| Kawasaki 2011[4] | 166 | Pediatric and adult | • New onset | • Aab titer<br>• Aab type<br>• Novel Aab/ epitope | Yes | • ZnT8A was more frequently observed in Japanese children than in adults at T1D diagnosis.<br>• ZNT8A concentration negatively associated with HLA-DR4.<br>• In children ZNT8A+ more likely to react to all 3 C-terminal variants of the ZnT8 protein. |
| Vermeulen 2011 (BDR)[5] | 1,416 | Pediatric and adult | • New onset | • Aab number<br>• Aab titer<br>• Aab type<br>• Novel Aab/ epitope | Yes | • IA-2bA and ZnT8A testing increased total multiple Aab+ individuals and certainty for immune-mediated disease if single IAA+, GAD+, or IA-2+.<br>• IA-2bA and ZnT8A associated with IA-2+ and younger age at diagnosis.<br>• IA-2bA (but not ZnT8A) associated positively with HLA-DQ8 and negatively with HLA-DQ2.<br>• Aab levels did not differ according to sex. |

Supplemental Table 2. Autoantibody features characterize heterogeneity at T1D diagnosis.

| | | | | | | |
|---|---|---|---|---|---|---|
| Gabbay 2012[6] | 60 | Pediatric | • New onset | • Aab number<br>• Aab titer<br>• Aab type | No | • IA-2 sera titers associated with a more inflammatory peripheral cytokine/chemokine profile than GAD.<br>• GAD titers negatively correlated with CXCL10 and CCL2 sera levels while IA-2 titers negatively correlated with IL-10 sera levels.<br>• No correlations between HLA genotype or PTPN22 SNP and GAD or IA-2 levels. |
| Long 2012 (BOX)[7] | 613 | Pediatric and adult | • New onset | • Aab number<br>• Aab type<br>• Aab timing | Yes | • During increased T1D incidence in the UK (1985-2002), IA-2A, IA-2bA and ZnT8 prevalence increased at diagnosis, but not IAA or GAD, suggesting a more intense humoral autoimmune response. |
| Ponsonby 2012[8] | 247 | Pediatric | • New onset | • Aab number<br>• Aab type | Yes | • IAA+ associated with younger age and red hair.<br>• GAD+ associated with lower birthweight and recent eczema.<br>• In <5yr, GAD+ associated with low recent or past sun exposure. |
| Trabucchi 2012[9] | 51 | Pediatric and adult | • New onset | • Aab titer<br>• Aab affinity<br>• Comparison of Aab assays | Yes | • Childhood-onset T1D patients presented lower proinsulin Aab concentrations and higher affinities than adults, suggesting a different etiopathogenic pathway. |
| Andersson 2013 (BDD)[10] | 3,165 | Pediatric | • New onset | • Aab number<br>• Aab type | Yes | • Adding ZnT8A to GAD, IA-2 and IAA increased frequency of Aab+ T1D from 90% to 93%.<br>• All 3 ZnT8A less frequent in children < 2yrs.<br>• Aab- patients more common under age < 2yrs and < 15 yrs.<br>• All 3 ZnT8A associated with HLA-DQA1-B1*X-0604 (DQ6.4) and DQA1-B1*03-0302 (DQ8) genotypes. |
| Lempainen 2013 (FPDR)[11] | 1,554 | Pediatric | • New onset | • Aab type | No | • INS rs689 SNP associated with IAA+ at diagnosis.<br>• IKZF4 rs1701704 C allele inversely associated with IAA+ at diagnosis.<br>• INS res689 and IKZF4 rs1701704 SNPs not associated with ICA+, GAD+, IA-2+, or ZnT8A+. |
| Long 2013 (BOX)[12] | 589 | Pediatric | • New onset | • Aab type<br>• Aab affinity | Yes | • HLA-A*24 associated with: lower IA-2+ and ZnT8A+, lower likelihood of IA-2 targeting tyrosine phosphatase domain of IA-2, and lower |

Supplemental Table 2. Autoantibody features characterize heterogeneity at T1D diagnosis.

| | | | | | | |
|---|---|---|---|---|---|---|
| | | | | • Novel Aab/epitope | | likelihood of ZnT8A targeting W325 polymorphic residue of ZnT8.<br>• No associations found with IAA or GAD. |
| Paschke 2013[13] | 344 | Adult | • New onset | • Aab number<br>• Aab type | Yes | • ICA+ and GAD+ more common in 18-35yrs vs. over 35yrs.<br>• Single AAb+ more common in over 35 yrs vs. 18-35.<br>• IA-2 most frequent single AAb in over 35yrs.<br>• Multiple AAb+ associated with younger age, lower fasting and stimulated c-peptide and shorter symptom duration at diagnosis. |
| Salonen 2013 (FPDR)[14] | 2,115 | Pediatric | • New onset | • Aab titer<br>• Aab type | Yes | • ZnT8+ associated with older age at diagnosis.<br>• DKA at diagnosis less common among ZnT8+ vs. ZnT8-.<br>• ZnT8+ decreased in DR3/DR4 heterozygotes vs. other DR combinations.<br>• Subjects with the neutral DR13-DQB1*0604 haplotype more frequently ZnT8+.<br>• T1D family hx not associated with ZnT8+ or levels. |
| Arif 2014[15] | 105 | Pediatric | • New onset | • Aab number<br>• Aab type | Yes | • Two distinct immune phenotypes were identified among individuals with new onset T1D and high risk siblings with multiple Aab: half with proinflammatory (IFN-gamma+, multiple Aab+) and half with partially regulated (IL-10+, pauci-aab+) phenotypes |
| Bollyky 2015[16] | 883 | Pediatric and adult | • New onset | • Aab type | Yes | • Adults less likely to be ICA+ or IA-2+ vs children <18yrs at T1D onset<br>• no difference in GAD+ between children and adults at T1D onset |
| Cedillo 2015[17] | 263 | Pediatric | • New onset | • Aab number<br>• Aab type | Yes | • No relationships between any measure of adiposity and Aab number, even with adjustment by age and/or HbA1c. |
| Kanatsuna 2015[18] | 1,039 | Pediatric | • New onset | • Aab type<br>• Novel Aab/epitope | No | • Aabs reactive with both insulin and INS-IGF2 at diagnosis were higher in patients than in controls irrespective of age at diagnosis.<br>• Specific INS-IGF2A levels did not correlate with other Aabs except for a week negative relationship with GAD. |

Supplemental Table 2. Autoantibody features characterize heterogeneity at T1D diagnosis.

| | | | | | | |
|---|---|---|---|---|---|---|
| | | | | | | • The risk of INS-IGF2A was increased among HLA-DQ2/8. |
| Maziarz 2015 (Swedish Childhood Diabetes & The Diabetes Incidence in Sweden Study Groups)[19] | 508 | Pediatric and adult | • New onset | • Aab type | No | • Increased risk of T1D by non-HLA genes was frequently modified by both Aabs and HLA-DQ.<br>• ERBB3 increased the risk of IA-2A+ T1D.<br>• Risk carrying G allele of 26471 associated with IAA- and IA-2- T1D.<br>• Risk carrying major allele A of IL2RA associated with IAA+ T1D and doubled the risk of T1D among subjects in the high-risk HLA group.<br>• INS genotype associated with IAA+ T1D but did not appear to be modified by HLA-DQ.<br>• OR of association between PTPN22 (CT+TT) and GAD+ T1D was higher compared to that for GAD- T1D.<br>• None of the other non-HLA genetic polymorphisms reported by T1D Genetics Consortium as significantly associated with T1D showed a relationship with T1D stratified by islet Aab. |
| Williams 2015 (BOX)[20] | 147 | Pediatric and adult | • New onset | • Aab type<br>• Novel Aab/epitope | Not available | • No difference between frequency of truncated GAD (96-585) vs. full antigen GAD (1-585) in children with new onset T1D. |
| Gómez-Díaz 2016[21] | 278 | Pediatric | • New onset | • Aab type | Yes | • In Mexican children with new onset T1D, HLA DRB1*04/DQA1*03/DQB1*03:02 haplotype associated with lower insulin, proinsulin, and C-peptide concentrations; IA-2+ impacted effect of this haplotype on lower proinsulin in a multivariable model. |
| Stoupa 2016[22] | 452 | Pediatric | • New onset | • Aab type | Yes | • Diabetes-associated Aabs and other immune markers did not statistically differ between ethnic groups (European Caucasian, Moghrabin Caucasian, Black African, and Mixed Origin)<br>• Positivity for ICA, IAA and IA-2 decreased with age.<br>• Positivity for GAD increased with age. |
| Bansal 2017[23] | 35 | Pediatric | • New onset, GAD+ only | • Novel Aab or epitope | Yes | • After adjustment for age, positive DPD epitope-specific GAD recognition was positively |

Supplemental Table 2. Autoantibody features characterize heterogeneity at T1D diagnosis.

| | | | | | | |
|---|---|---|---|---|---|---|
| | | | | | | associated with higher C-peptide levels at T1D diagnosis.<br>• In children over 10 yrs, sex-adjusted BMI percentile correlated with recognition of the DPD-defined epitope.<br>• No independent associations of DPD-defined epitope with sex, race/ethnicity, DKA, HbA1c, HLA DR3-DQ2/DR4-DQ8 or Aab number. |
| Spanier 2017[24] | 63 | Pediatric and adult | • New onset | • Aab titer<br>• Aab type | Not available | • HLA-DQ8+ patients with T1D have increased tetramer+CD4+T cells compared with HLA-matched control subjects without diabetes.<br>• Shorter disease duration was associated with higher frequencies of insulin-reactive CD4+T cells<br>• Insulin tetramer+ effector memory cells is correlated with IAA titers.<br>• One of 4 control subjects with tetramer+ cells was FDR who had insulin-specific cells with an effector memory phenotype, potentially representing an early marker of T-cell autoimmunity. |
| Viisanen 2017 (DIPP)[25] | 276 | Pediatric | • Multiple Aab+<br>• New onset | • Aab number<br>• Aab type | No | • Frequency of CXCR5+PD-1+ICOS+ activated Tfh cells is increased in children with new onset T1D and multiple Aab+ children with impaired glucose tolerance.<br>• No alterations in circulating B cell compartments before or after T1D onset. |
| Alyafei 2018[26] | 424 | Pediatric | • New onset | • Aab number<br>• Aab type | Yes | • Prevalence of GAD and IAA did not differ between familial or non-familial T1D<br>• ICA more prevalent in familial vs non-familial T1D<br>• Prevalence of being positive for all three (ICA, GAD, IAA) did not differ between familial and non-familial T1D. |
| Bravis 2018[27] | 1,778 | Pediatric and adult | • New onset | • Aab number<br>• Aab type | Yes | • 85% of participants with a clinical T1D diagnosis were GADA, IA2A, or ZnT8A Aab+.<br>• Presenting symptoms and DKA frequency similar between Aab+ and Aab-. |

Supplemental Table 2. Autoantibody features characterize heterogeneity at T1D diagnosis.

| | | | | | | |
|---|---|---|---|---|---|---|
| | | | | | | • Aab+ less common with increasing age, in males, and in non-white compared with white race.<br>• Aab- adults with higher BMI, more likely to have parent with diabetes.<br>• Aab- less likely to have other autoimmune disease. |
| Ilonen 2018 (FPDR)[28] | 1,028 | Pediatric | • New onset | • Aab type<br>• Aab timing | Yes | • At diagnosis, IA-2 was most common, followed by IAA, ZnT8, and GAD<br>• Aab combinations at diagnosis stratified into groups with either IAA or GAD first and were associated with specific risk genotypes (*INS* for IAA first and *IKZF4-ERBB3* for GAD first) in children diagnosed <10 yrs |
| Niechciał 2018[29] | 367 | Pediatric and adult | • New onset | • Aab number<br>• Aab titer<br>• Aab type | Yes | • Children mostly positive for 2 (37.8%) and 3 (49.5%) Aabs, vs. adults for 1 (32.2%) and 2 (30.7%).<br>• Most frequently detected Aabs in youth were ZnT8A (81.1%) and IA-2 (80.7%), vs. GAD in adults (74.8%).<br>• ZnT8 titers higher in children, but adults with higher GAD and IA-2 titers.<br>• ZnT8A+ and IA-2+ reported mostly in individuals with DKA.<br>• Number of Aabs correlated with severity of DKA.<br>• ZnT8 associated with a greater risk of DKA independent of gender, age group and Aab number. |
| Niechcial 2018[30] | 735 | Pediatric | • New onset | • Aab number<br>• Aab titer<br>• Aab type | Yes | • No associations found between other Aabs and DKA at diagnosis.<br>• Higher occurrence and titers of ZnT8 were reported in those diagnosed with DKA. |
| Redondo 2018[31] | 810 | Pediatric and adult | • New onset | • Aab number<br>• Aab type | Yes | • Persons >12 yrs with rs45065659 and 7901695 TCF7L2 variants were more likely to have single Aab+, higher C-pep AUC and lower mean glucose at diagnosis. |
| Turtinen 2018 (FPDR)[32] | 4,993 | Pediatric | • New onset | • Aab number<br>• Aab titer | No | • Boys were more often IAA+, IA-2+ and ZnT8+<br>• Girls had higher frequency of GAD+ Aabs |

Supplemental Table 2. Autoantibody features characterize heterogeneity at T1D diagnosis.

| | | | | | | |
|---|---|---|---|---|---|---|
| | | | | • Aab type | | |
| Sales Luis 2019[33] | 137 | Pediatric | • New onset | • Aab number<br>• Aab type | Yes | • Children diagnosed with T1D at <5yrs were more commonly positive for three Aabs than children diagnosed >5yrs.<br>• Children diagnosed with T1D at <5yrs were more commonly IAA+ |
| Turtinen 2019 (FPDR)[34] | 4993 | Pediatric | • New onset | • Aab number<br>• Aab timing | No | • Children with familiar T1D (at least one affected mother, father, or sibling) were more likely to test negative for all Aabs and were more frequently IAA+ |
| Vicinanza 2019[35] | 532 | Pediatric | • New onset | • Aab number<br>• Aab type | Yes | • Only single positive IAA had significantly higher frequencies in children with DKA compared to children without DKA at diagnosis.<br>• No solitary positive AAb were significantly associated to any degree of DKA.<br>• No significant differences in the prevalence of any multiple AAb positivity at diagnosis neither concerning the occurrence of DKA nor its severity. |
| Luo 2020[36] | 751 | Pediatric and adult | • New onset | • Aab number<br>• Aab titer<br>• Aab type | Yes | • Children with acute onset T1D showed higher prevalence of IA-2+, ZnT8+, and multiple autoantibodies than adults with acute onset T1D<br>• There was an overall decreasing frequency of IA-2+ and multiple Aab+ with increasing age.<br>• The highest frequency of IA-2 and multiple Aab+ were in patients <10 yrs.<br>• Children who were 10 to 19.9 yrs had the highest prevalence of GAD.<br>• In children with acute onset T1D, DR3 is related to ZnT8, and DR3/DR9 is related to IA-2 and multiple Aab+, but not adults with acute onset T1D. |
| Redondo 2020 (TrialNet)[37] | 786 | Pediatric and adult | • New onset | • Aab number<br>• Aab type | Yes | • Single Aab+ at dx observed in 15.1% of participants.<br>• Single Aab+ associated with older age, higher C-peptide measures, higher HOMA-IR, and lower T1D Index60 measure. |

Supplemental Table 2. Autoantibody features characterize heterogeneity at T1D diagnosis.

| | | | | | | |
|---|---|---|---|---|---|---|
| | | | | | | • After BMI adjustment, associations with age and 2-hour C-peptide no longer significant. |
| Jia 2021[38] | 302 | Pediatric | • New onset | • Comparison of Aab assays | Not available | • No difference in ZnT8 positivity for ECL or RBA assays in new onset T1D (62%). |
| Nieto 2021[39] | 711 | Pediatric | • New onset | • Aab number<br>• Aab type | Yes | • In multivariable analysis, IAA+ associated with younger age and lower HbA1c; but not Tanner stage, GAD+ or Aab+.<br>• IAA titers associated with younger age, more DKA, and higher tTGA levels.<br>• GAD+ associated with female sex, negatively associated with + tTGA titers, but not with age, IAA+, IA-2+, Aab number, or thyroid autoimmunity.<br>• GAD titers associated with female sex, racial minority status and TPO+.<br>• IA-2+ not associated with any variables tested. IA-2 titers associated with older age and not being African American. |
| Williams 2021[40] | 401 | Pediatric and adult | • New onset | • Aab number<br>• Aab titer<br>• Aab type | Yes | • Detectable residual C-peptide post-diagnosis was best predicted by a combined model with incorporation of duration, age at onset, GRS, and titers for GAD, IA-2, and ZnT8.<br>• GAD titer was directly related to the odds of C-peptide detection.<br>• Subjects with low or undetectable titers of IA-2 and ZnT8 at diagnosis may experience a slower decline in functional beta-cell mass. |
| Fakhfakh 2022[41] | 156 | Pediatric | • New onset | • Aab type | Yes | • ZnT8A levels correlated weakly with age at dx. |
| Jacobsen 2022 (TEDDY)[42] | 379 | Pediatric | • New onset | • Aab number<br>• Aab type<br>• Age at Aab seroconversion<br>• Order of Aab seroconversion | Yes | • Younger children had fewer Aabs with more symptoms at diagnosis.<br>• Incidence of T1D was highest for those presenting with multiple Aabs as first appearance of Aabs as compared with either IAA+ or GAD+ first.<br>• No significant difference in IAA or GAD first after adjusting for age at seroconversion.<br>• Children who develop Aabs and progress to T1D early in life have less functional beta-cell mass and higher rates of DKA at diagnosis. |

Supplemental Table 2. Autoantibody features characterize heterogeneity at T1D diagnosis.

| | | | | | | |
|---|---|---|---|---|---|---|
| Parviainen 2022 (FPDR)[43] | 6,015 | Pediatric | • New onset | • Aab number<br>• Aab type<br>• Order of Aab seroconversion | Yes | • Children diagnosed at <7yrs had stronger familial clustering and HLA-DR/DQ-conferred risk, higher number of Aabs at diagnosis, and higher frequency of IAA+ when compared with older children.<br>• Children diagnosed at or over 13 yrs years were more often male, had weaker familial clustering and HLA conferred genetic risk<br>• Children diagnosed at or over 13 yrs were more frequently GAD+, had longer duration of symptoms before diagnosis, and a higher frequency of diabetic ketoacidosis.<br>• ICA titers did not differ with age at diagnosis. |
| Taka 2022 (FPDR)[44] | 5,798 | Pediatric | New onset | • Aab number<br>• Aab titer<br>• Aab type | Yes | • Frequency of ICA, IAA, IA-2+ was higher in subjects with high/moderate risk HLA genotypes<br>• Frequency of GAD+ was higher in subjects with other HLA genotypes<br>• No significant difference in ZnT8+ was found between the groups with high/moderate risk HLA vs other HLA<br>• In decreasing order of detection rate across both groups, ICA was the most frequently detected followed by IA-2, GAD, and ZnT8 with similar rates, and IAA.<br>• High/moderate risk HLA subjects had more participants with multiple Aabs whereas Aab- was more frequent in other HLA subjects.<br>• Levels of IAA and IA-2 were higher in high/moderate risk HLA whereas ZnT8 levels were higher in other HLA subjects.<br>• No significant differences in ICA or GAD levels were found between the two groups. |

*Participant groups were considered new onset if within 12 months of T1D diagnosis

Abbreviations: Aab (autoantibody), ADBW-END (Arbeitsgemeinschaft Diabetologie Baden-Württemberg - Erhebung Neu entdeckter Typ 1 Diabetiker Study group), BDD (Better Diabetes Diagnosis), BDR (Belgian Diabetes Registry), DSBD (Danish Study Group of Childhood Diabetes), BOX (Bart’s Oxford Family Study), FDR (first degree relative), FPDR (Finnish Pediatric Diabetes Register), GAD (glutamic acid decarboxylase

Supplemental Table 2. Autoantibody features characterize heterogeneity at T1D diagnosis.

antibody), HOMA-IR (homeostatic model assessment of insulin resistance), IA-2 (islet antigen-2 antibody), IAA (insulin autoantibody), ICA (islet cell autoantibody), ZnT8 (zinc transporter antibody)

Supplemental Table 3. Autoantibody features characterize progression after T1D diagnosis.

| Study | n | Age group | Population Studied* | Autoantibody Feature Assessed | Age impact? | Findings |
|---|---|---|---|---|---|---|
| Hameed 2011[1] | 367 | Pediatric | • New onset | • Timing of Aab development | No | • Persistent Aab- status at and after diagnosis associated with preserved residual C-peptide.<br>• Aab- children testing negative for monogenic diabetes exhibited high frequency of diabetogenic HLA. |
| Nielsen 2011 (Hvidovre Study Group on Childhood Diabetes)[2] | 257 | Pediatric | • New onset | • Aab type<br>• Novel Aab or epitope | Yes | • ZnT8R epitope Aab+ was more frequent in those >5yrs at diagnosis while ZnT8W epitope Aab+ was not age-related<br>• No relationship between ZnT8A+, ICA+, IA-2+ at 1 month post-diagnosis and residual C-peptide at 12 months post-diagnosis.<br>• IAA+ or GAD+ at 1 month post-diagnosis associated with lower 12 month stimulated C-peptide.<br>• GAD+ associated with 12 month HbA1c. |
| Andersen 2012 (Danish Remission Phase Study)[3] | 129 | Pediatric | • New onset | • Aab type<br>• Aab titer<br>• Novel Aab or epitope | Yes | • All ZnT8A variant titers decreased over 12 months post-diagnosis.<br>• Higher arginine variant of ZnT8 Aab associated with higher C-peptide over 12 months post-diagnosis.<br>• Positive correlation between all three ZnT8 Aab variants and IA-2 titers over 12 months post-diagnosis (but not GAD or IAA). |
| Sorensen 2012 (Danish Registry for Childhood Diabetes)[4] | 260 | Pediatric | • New onset | • Aab number<br>• Aab titer<br>• Aab type | Yes | • Reductions in IA-2, ZnT8W, or ZnT8Q (but not ZnT8R or GAD) titers over 3-6 years post-diagnosis associated with higher likelihood of detectable C-peptide. |
| Chao 2013[5] | 247 | Pediatric and adult | • New onset | • Aab type | No | • GAD+ more common than IA-2+ at diagnosis in Chinese patients with acute onset T1D (56.3% vs 32.8%).<br>• Most patients remained GAD+ or IA-2+ during follow-up.<br>• C-peptide values were higher in GAD- or IA-2- patients compared to GAD+ or IA-2+ at diagnosis, independently of whether Aab positivity persisted over time or not. |

Supplemental Table 3. Autoantibody features characterize progression after T1D diagnosis.

| | | | | | | |
|---|---|---|---|---|---|---|
| Ludvigsson 2013 (BDD)[6] | 4017 | Pediatric | • New onset | • Aab type | Yes | • IAA+ was associated with more rapid post-diagnosis C-peptide loss when controlling for age<br>• No relation to GAD or IA-2 positivity was detected. |
| Pecheur 2014[7] | 242 | Pediatric | • New onset | • Aab number<br>• Aab type | Yes | • Partial remission duration was decreased in single Aab+ versus both GAD+ and IA-2+. |
| Stoupa 2016[8] | 452 | Pediatric | • New onset | • Aab type | Yes | • Mean C-peptide at 2 years post-diagnosis was correlated with absence of ICA or IAA at diagnosis in European ethnic groups (European Caucasian, Moghrabin Caucasian, Black African, and Mixed Origin). |
| Marino 2017[9] | 204 | Pediatric | • New onset | • Aab number | Yes | • Higher Aab+ number at diagnosis associated with lower rates of partial remission. |
| Camilo 2020[10] | 51 | Pediatric and adult | • New onset | • Aab number<br>• Aab type | Yes | • No significant difference in partial remission rates in GAD+ compared to IA-2+ Brazilian children.<br>• HLA DRB1*0301-DQB1*0201 associated with lower IA-2+ and higher remission rates. |
| Steck 2021 (TEDDY)[11] | 113 | Pediatric | • New onset | • Aab number<br>• Aab type | Yes | • Higher Aab+ number at diagnosis associated with higher rate of C-peptide loss in univariate analysis.<br>• IA-2+ or ZnT8+ at diagnosis associated with higher rate of C-peptide loss in univariate analysis.<br>• Relationships no longer statistically significant in multivariate analysis including age, sex, and weight z-score. |

*Participant groups were considered new onset if they were within 12 months of T1D diagnosis.

Abbreviations: Aab (autoantibody), BDD (Better Diabetes Diagnosis), FDR (first degree relative), GAD (glutamic acid decarboxylase antibody), IA-2 (islet antigen-2 antibody), IAA (insulin autoantibody), ICA (islet cell autoantibody), TEDDY (The Environmental Determinants of Diabetes in the Young), ZnT8 (zinc transporter antibody)

Supplemental Table 3. Autoantibody features characterize progression after T1D diagnosis.

Supplemental Table 4. Autoantibody features characterize heterogeneity in responses to disease modifying therapy.

| Study | n | Age group | Population Studied* | Autoantibody Feature Assessed | Age impact? | Findings |
|---|---|---|---|---|---|---|
| Christie 2002[1] | 97 | Pediatric and adult | • New onset | • Aab type | No | • Cyclosporin had no significant effect on frequency of IA-2+ Aab.<br>• Cyclosporin reduced insulin requirements and increased C-peptide secretion in IA-2- participants.<br>• IA-2+, GAD- participants were most resistant to cyclosporin.<br>• No differential effects were observed for partitioning by ICA+ or IAA+. |
| Gale 2004 (ENDIT)[2] | 552 | Pediatric and adult | • FDR (ICA+) | • Aab number<br>• Aab type | No | • No difference in time to diabetes noted between nicotinamide and placebo groups in the Cox proportional hazard estimate when adjusted for Aab number.<br>• No evidence of a nicotinamide treatment effect in groups divided by Aab status. |
| Skyler 2005 (DPT-1)[3] | 372 | Pediatric and adult | • FDR<br>• Single Aab+<br>• Multiple Aab+ | • Aab titer<br>• Aab type | Not available | • Oral insulin did not delay or prevent type 1 diabetes progression in ICA+ and IAA+ relatives. |
| Näntö-Salonen 2008[4] | 264 | Pediatric | • High genetic<br>• FDR<br>• Multiple Aab+ | • Aab number<br>• Aab titer<br>• Aab type | No | • Aab features did not impact ability of nasal insulin to delay or prevent T1D. |
| Pescovitz 2009 (TrialNet)[5] | 87 | Pediatric and adult | • New onset | • Aab number<br>• Aab titer<br>• Aab type | No | • No significant differential treatment effect of rituximab among subgroups based Aabs. |
| Wherrett 2011 (TrialNet)[6] | 145 | Pediatric and adult | • New onset (GAD+) | • Aab type | No | • Subcutaneous GAD-alum did not preserve insulin secretion in GAD+ participants with recently diagnosed T1D. |
| Yu 2011 (TrialNet)[7] | 87 | Pediatric and adult | • New onset | • Aab number<br>• Aab titer<br>• Aab type | Not available | • Rituximab suppressed IAAs compared with placebo but had smaller effect on GAD, IA-2, and ZnT8 Aabs at 1 year.<br>• 40% IAA+ individuals treated with rituximab became IAA- (none became negative with placebo). |

Supplemental Table 4. Autoantibody features characterize heterogeneity in responses to disease modifying therapy.

| | | | | | | |
|---|---|---|---|---|---|---|
| | | | | | | • Rituximab within 50 days of T1D diagnosis led to marked suppression of IAA for 1-3 yrs.<br>• IAA levels were lower for individuals who maintained C-peptide levels during 1st year after diagnosis but this was independent of rituximab treatment. |
| Ludvigsson 2012 (Diamyd)[8] | 334 | Pediatric and adult | • New onset | • Aab titer<br>• Aab type | No | • In GAD+ new onset T1D, treatment with alum-formulated GAD65 did not reduce loss of stimulated C-peptide compared to placebo.<br>• Stratification based on baseline GAD titer did not impact treatment response. |
| Herold 2013 (ITN-AbATE)[9] | 77 | Pediatric and adult | • New onset | • Aab type | No | • Significant reduction in the titer of ZnT8 (but not IA-2, IAA, or GAD) in teplizumab-treated participants after 1 year but not after 2 years.<br>• Baseline individual Aab positivity did not predictor response to teplizumab. |
| Aronson 2014 (DEFEND-1)[10] | 272 | Pediatric and adult | • New onset | • Aab number<br>• Aab type | No | • No significant impact of GAD or IA-2 positivity or Aab number on Otelixizumab treatment effect. |
| Demeester 2015[11] | 80 | Pediatric and adult | • New onset | • Aab number<br>• Aab type | No | • Higher IAA levels were associated with better preservation of beta cell function and lower insulin with anti-CD3 treatment.<br>• In multivariate analysis, IAA or the interaction of IAA and C-peptide independently predicted outcome together with treatment.<br>• During follow-up, anti-CD3 responders (i.e., IAA+ participants with preserved beta cell function) showed a less pronounced insulin-induced rise in IAA and lower insulin needs.<br>• GAD, IA-2, and ZnT8 Aab levels were not influenced by anti-CD3, and their changes showed no relationship with outcomes. |
| Krischer 2017 (TrialNet)[12] | 560 | Pediatric and adult | • FDR<br>• Second degree relative<br>• Multiple Aab+ | • Aab number<br>• Aab titer<br>• Aab type | Not available | • Among multiple Aab+ relatives with a high IAA titer, 7.5 mg/d oral insulin, did not delay or prevent T1D development vs. placebo. |

Supplemental Table 4. Autoantibody features characterize heterogeneity in responses to disease modifying therapy.

| | | | | | | |
|---|---|---|---|---|---|---|
| | | | • Other: third degree relative | | | |
| Herold 2019 (TrialNet)[13] | 76 | Pediatric and adult | • Multiple Aab+ | • Aab type | No | • Response to teplizumab was greater if ZnT8, GAD, or IAA were positive, and if IA-2 or IAA were negative. |

*Participant groups were considered new onset if they were within 12 months of T1D diagnosis.

Abbreviations: Aab (autoantibody), AbATE (Autoimmunity-Blocking Antibody for Tolerance in Recently Diagnosed Type 1 Diabetes), ENDIT (European Nicotinamide Diabetes Intervention Trial), DEFEND-1 (Durable Response Therapy Evaluation for Early or New-Onset Type 1 Diabetes), DPT-1 (Diabetes Prevention Trial Type 1), FDR (first degree relative), GAD (glutamic acid decarboxylase antibody), IA-2 (islet antigen-2 antibody), IAA (insulin autoantibody), ICA (islet cell autoantibody), ITN (Immune Tolerance Network), ZnT8 (zinc transporter antibody)
